# Emergency care interventions for pediatric severe acute respiratory infections in low- and middle-income countries: a systematic review and narrative synthesis

**DOI:** 10.1101/2023.01.19.23284769

**Authors:** Pryanka Relan, Stephanie Chow Garbern, Gerard O’Reilly, Corey B Bills, Megan Schultz, Sean Kivlehan, Indi Trehan, Torben K Becker

## Abstract

**Background:** Severe acute respiratory infections (SARIs) are the leading cause of pediatric death globally, particularly in low- and middle-income countries (LMICs). Given the potential rapid clinical decompensation and high mortality rate from SARIs, interventions that facilitate the early care of patients are critical to improving patient outcomes. The aim of this systematic review was to evaluate the impact of emergency care interventions on improving clinical outcomes of pediatric patients with SARIs in LMICs.

**Methods:** PubMed, Global Health, and Global Index Medicus electronic databases were searched to identify peer-reviewed clinical trials or studies with comparator groups published before November 2020. All studies which evaluated acute and emergency care interventions on clinical outcomes for children (29 days to 19 years) with SARIs conducted in LMICs were included. Given the heterogeneity of interventions and outcomes, a narrative synthesis was performed. Bias was assessed using the Risk of Bias 2 and Risk of Bias in Non-Randomized Studies of Interventions.

**Results:** 20,583 studies were screened, with 99 meeting criteria for final inclusion. The most common conditions studied were pneumonia or acute lower respiratory infection (61.6%) and bronchiolitis (29.3%). The majority of studies evaluated medications (80.8%), followed by respiratory support (14.1%) and supportive care (5%). The strongest evidence of benefit was found for respiratory support interventions such as improved medical oxygen systems to decrease risk of death. Results were notably inconclusive on the utility of continuus positive airway pressure (CPAP). Mixed results were found for interventions for bronchiolitis, although possible benefit was found for hypertonic nebulized saline to decrease hospital length of stay. Early use of adjuvant treatments such as Vitamin A, D, and zinc for pneumonia and bronchiolitis did not appear to have convincing evidence of benefit on clinical outcomes.

**Conclusions:** While the burden of SARI in pediatric populations is high, few emergency care (EC) interventions have high quality evidence for benefit on clinical outcomes in LMICs. Respiratory support interventions have the strongest evidence for decreasing hospital length of stay and mortality and improvement of clinical status. Further research on the use of CPAP in diverse settings and populations is needed. A stronger evidence base for EC interventions for children with SARI, including metrics on the timing of interventions, is greatly needed.

## INTRODUCTION

Acute respiratory infections (ARIs) are the leading cause of pediatric morbidity and mortality in low- and middle-income countries (LMICs), with pneumonia being the leading infectious cause of death in children under five years globally, causing over 800,000 deaths in 2019.^1,2^ARIs encompass a broad range of conditions from a simple “cold” or upper respiratory infection to bronchiolitis, pneumonia, respiratory involvement of measles and complications such as acute respiratory distress syndrome (ARDS)^3,4^. The World Health Organization (WHO) definition of Severe Acute Respiratory Infection (SARI) was developed in 2011 and updated in 2017 with the aim of standardizing surveillance for influenza-like illness (ILI) and was defined as “an acute respiratory infection with cough and fever which requires hospitalization.”^5^

SARIs can be caused by a variety of pathogens, including viruses, bacteria, and parasites. Certain viral etiologies of SARI pose pandemic potential, such as pandemic influenza A (H1N1/09), and severe acute respiratory syndrome coronavirus 2 (SARS-CoV-2). Children in LMICs are among the most vulnerable to having poor outcomes from SARI due to inadequate immunization coverage, underlying conditions such as malnutrition, as well as limited access to health care resources such as medical oxygen, ventilators and relevant therapeutics. Emergency care (EC) interventions (i.e., those that provide or facilitate the early care of patients) are pivotal in improving outcomes for children with SARI.

Despite advances in characterizing the etiologies, incidence, and factors contributing to SARIs, identifying which EC interventions are most effective in improving clinical outcomes in LMICs is critical to ensuring that limited available resources can be optimally targeted towards feasible and effective interventions. This systematic review’s aim was to evaluate the impact of EC interventions on improving clinical outcomes of children with SARIs in LMICs.

## METHODS

This review was conducted according to the Preferred Reporting Items for Systematic Reviews and Meta-analyses (PRISMA) guidelines and in collaboration with the Global Emergency Medicine Literature Review (GEMLR) groups^6^ and registered on PROSPERO (CRD42020216117). As only published, de-identified data were used, this study was exempt from institutional review board approval^7^. A full description of the systematic review methods has been previously published in the concurrently conducted adult-focused review and are summarized below.^7^

### Search Strategy and Inclusion Criteria

PubMed, Global Health, and Global Index Medicus databases were searched November 2020 to January 2021. Eligible studies included all randomized controlled trials (RCTs) and observational studies with a control group that evaluated short-term clinical outcomes and included each of three major search themes: SARI, emergency care interventions, and LMICs as defined by the 2020 World Bank Classification. Two independent reviewers screened each title and abstract using Covidence, with discrepancies resolved by a third reviewer; the same procedure was followed for full-text screening. Both pediatric and adult articles were included during initial screening, and later separated into pediatric (29 days to 19 years, as per WHO definitions) and adult-focused reviews, given the variations in SARI epidemiology and management between adults and children. Studies that focused on neonates (<28 days) only were excluded. Studies were screened based on the WHO SARI case definition criteria (i.e., regardless of whether the term “SARI” itself was used) due to a lack of consistent implementation of SARI surveillance across all LMIC contexts^7^. EC interventions were defined as relevant interventions initiated in the early period of care (within ∼24 hours) without restrictions on setting. Only studies which evaluated patient-centric clinical outcomes such as mortality, need for intensive care unit (ICU) admission or mechanical ventilation, hospital length-of-stay (LOS), clinical severity, symptom duration or severity, were included (Appendix 1).

### Assessment of Risk of Bias

Two authors independently assessed the included studies for risk of bias using the Cochrane Risk of Bias 2 (RoB 2) tool for randomized trials and the Risk of Bias in Non-Randomized Studies (ROBINS-I) tool for observational studies; discrepancies were resolved by a third author. Risk of bias plots were created using *robvis* in R^8^.

### Data Extraction and Analysis

Two authors independently evaluated each article and extracted the data on a standardized form using Covidence, with discrepancies resolved by a third author (Appendix 2). Due to the heterogeneity of interventions, settings, and outcomes measured, requisite criteria for formal meta-analyses were not met, and therefore, a qualitative analysis and narrative synthesis was undertaken.

## RESULTS

The initial literature search yielded 25,180 studies, with 4,957 duplicates removed. After screening titles, abstracts, and full-text articles, 99 studies met inclusion criteria for further analysis (Figure 1). Pneumonia was the most common condition studied (52.5%), followed by bronchiolitis (29.3%, Table 1). Medications were the most common intervention type (80.8%) followed by respiratory support (14.1%). Most studies were conducted in general hospital/pediatric wards (72.7%), while only 18.1% of studies enrolled patients in emergency wards. Further characteristics of the studies included are shown in Table 1. Given the substantial number of studies, the results are presented by type of intervention, followed by the disease/condition studied (Table 2). Results obtained from the risk of bias assessment for RCTs and for observational studies are summarized in Figure 2 and 4, with individual risk of bias assessment for each study shown in Figures 3 and 5.

**Table 1.** Characteristics of studies (n=99).

|  | **Number of studies (%)**<br>**(N=99)** |
| --- | --- |
| **Type of study** |  |
| Randomized controlled trial | 91 (91.9) |
| Observational, prospective | 4 (4.0) |
| Observational, retrospective | 3 (3.0) |
| Non-randomized trial | 1 (1.0) |
| **World Bank income group** |  |
| Lower middle-income only | 60 (60.6) |
| Upper middle-income only | 23 (23.2) |
| Low-income only | 12 (12.1) |
| Mixed: LIC, LMIC, UMIC | 4 (4.0) |
| **World Health Organization region** |  |
| Southeast Asian | 25 (25.3) |
| Eastern Mediterranean | 24 (24.2) |
| African | 21 (21.2) |
| American | 14 (14.1) |
| Western Pacific | 12 (12.1) |
| Mixed | 2 (2.0) |
| European | 1 (1.0) |
| **Disease/condition** |  |
| Pneumonia | 52 (52.5) |
| Bronchiolitis | 29 (29.3) |
| Acute lower respiratory tract infection | 9 (9.1) |
| Undifferentiated acute respiratory distress | 5 (5.1) |
| Croup | 2 (2.0) |
| Influenza | 1 (1.0) |
| Severe Acute Respiratory Illness | 1 (1.0) |
| **Intervention** |  |
| Medications | 80 (80.8) |
| Respiratory support | 14 (14.1) |
| Supportive care | 5 (5.0) |
| **Location where study took place** |  |
| Emergency department | 18 (18.1) |
| Hospital ward | 72 (72.7) |
| Intensive care unit | 3 (3.0) |
| Outpatient / Other / Not specified | 6 (6.0) |

**Table 2.**
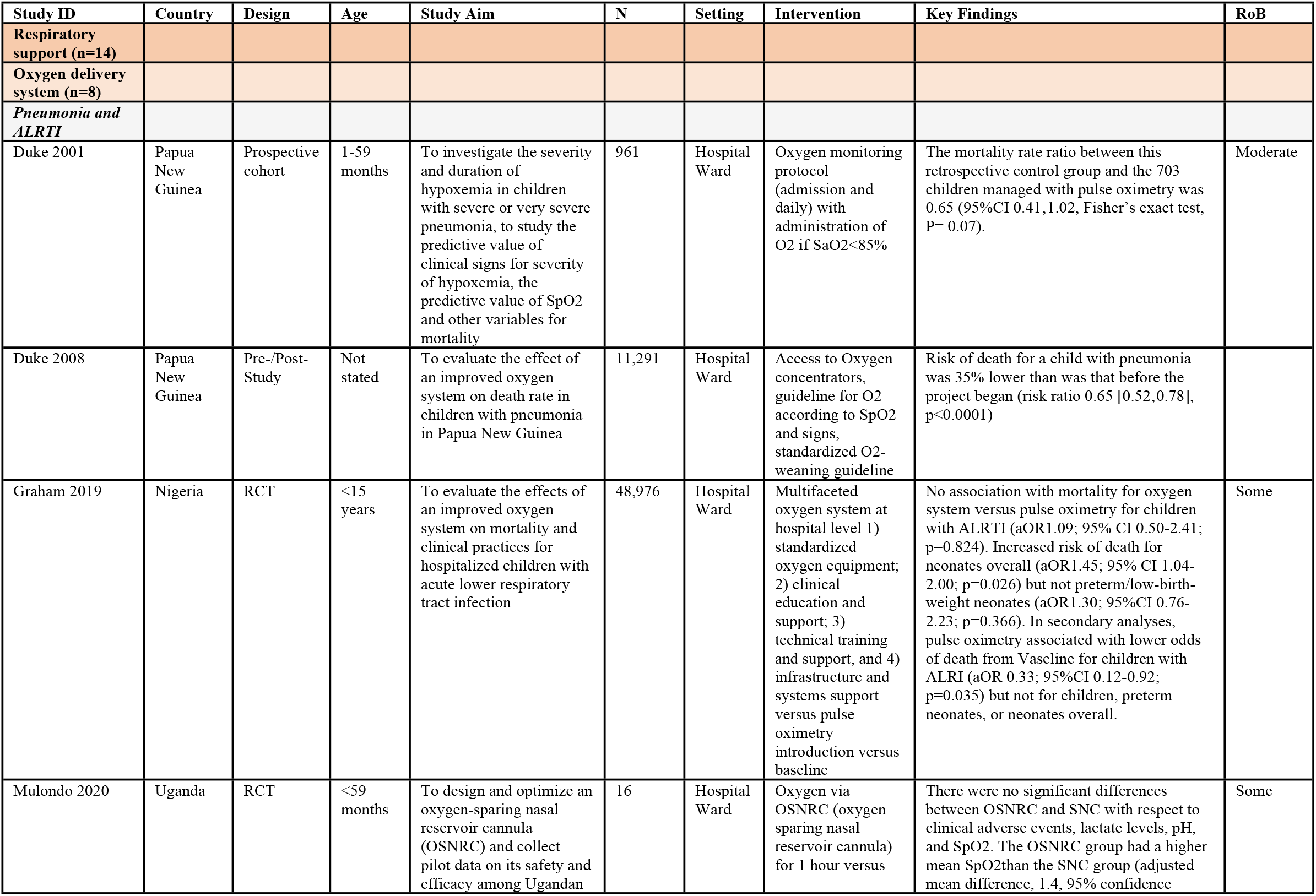
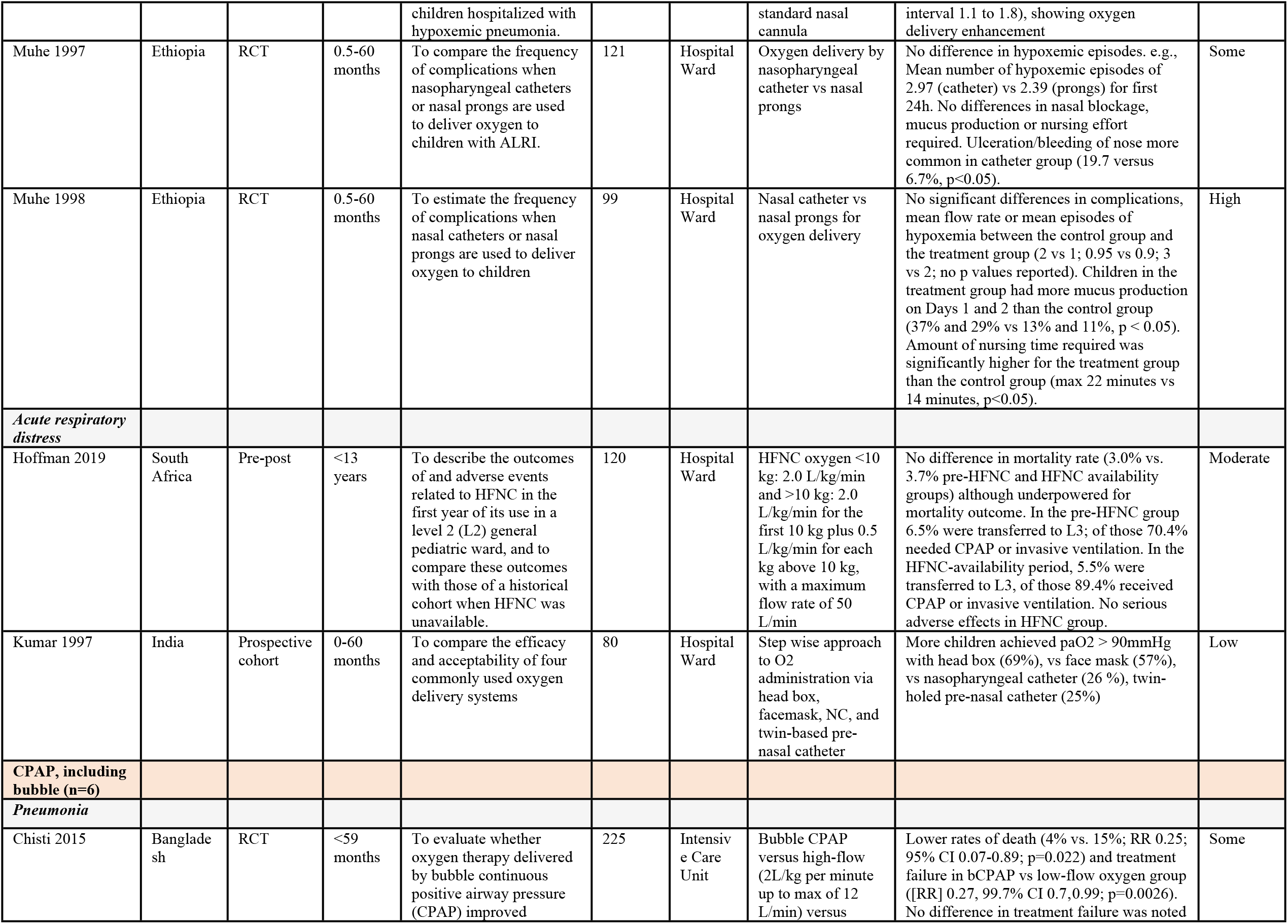
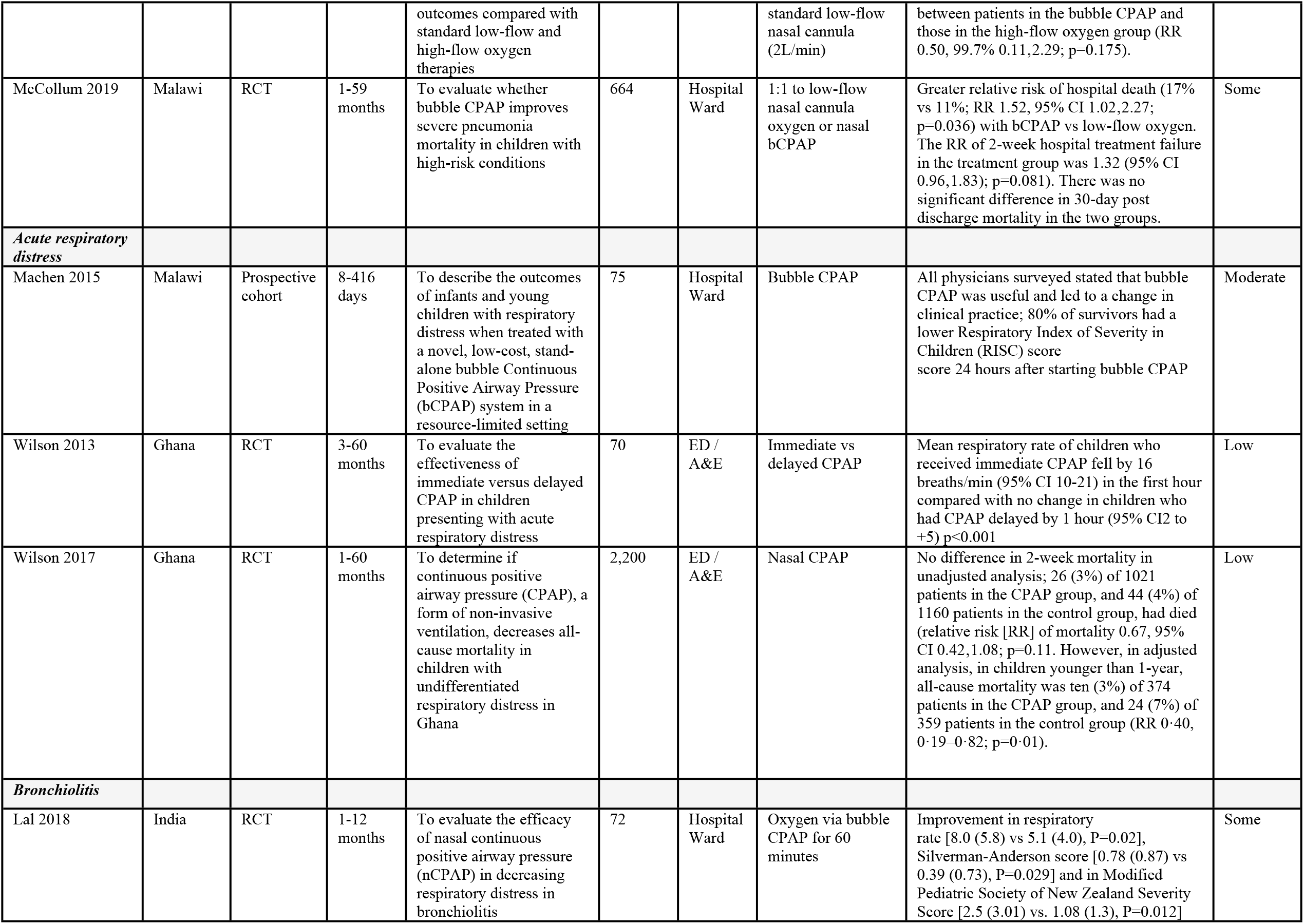
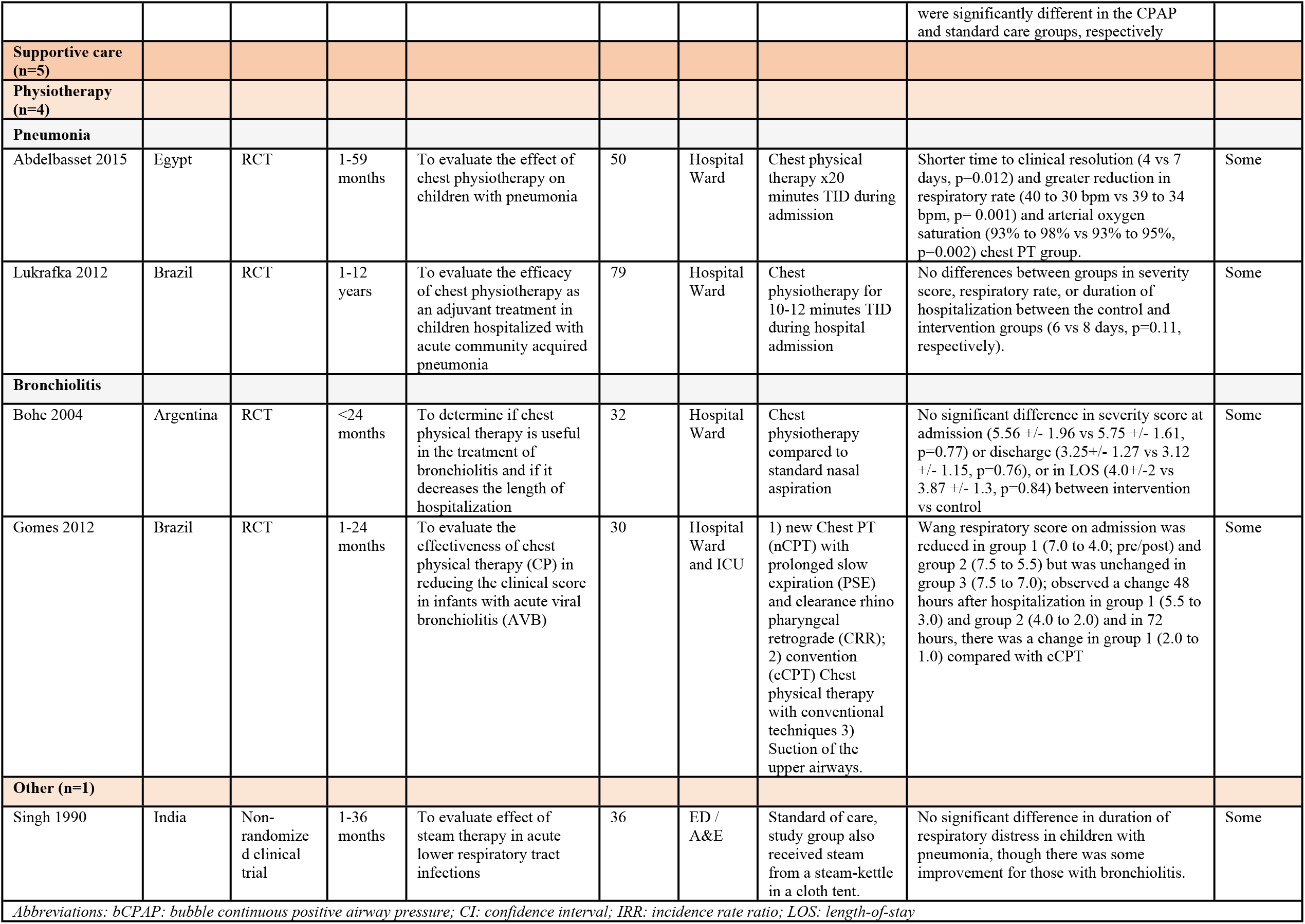
Summary of Findings – Respiratory Support and Supportive Care.

**Table 3.**
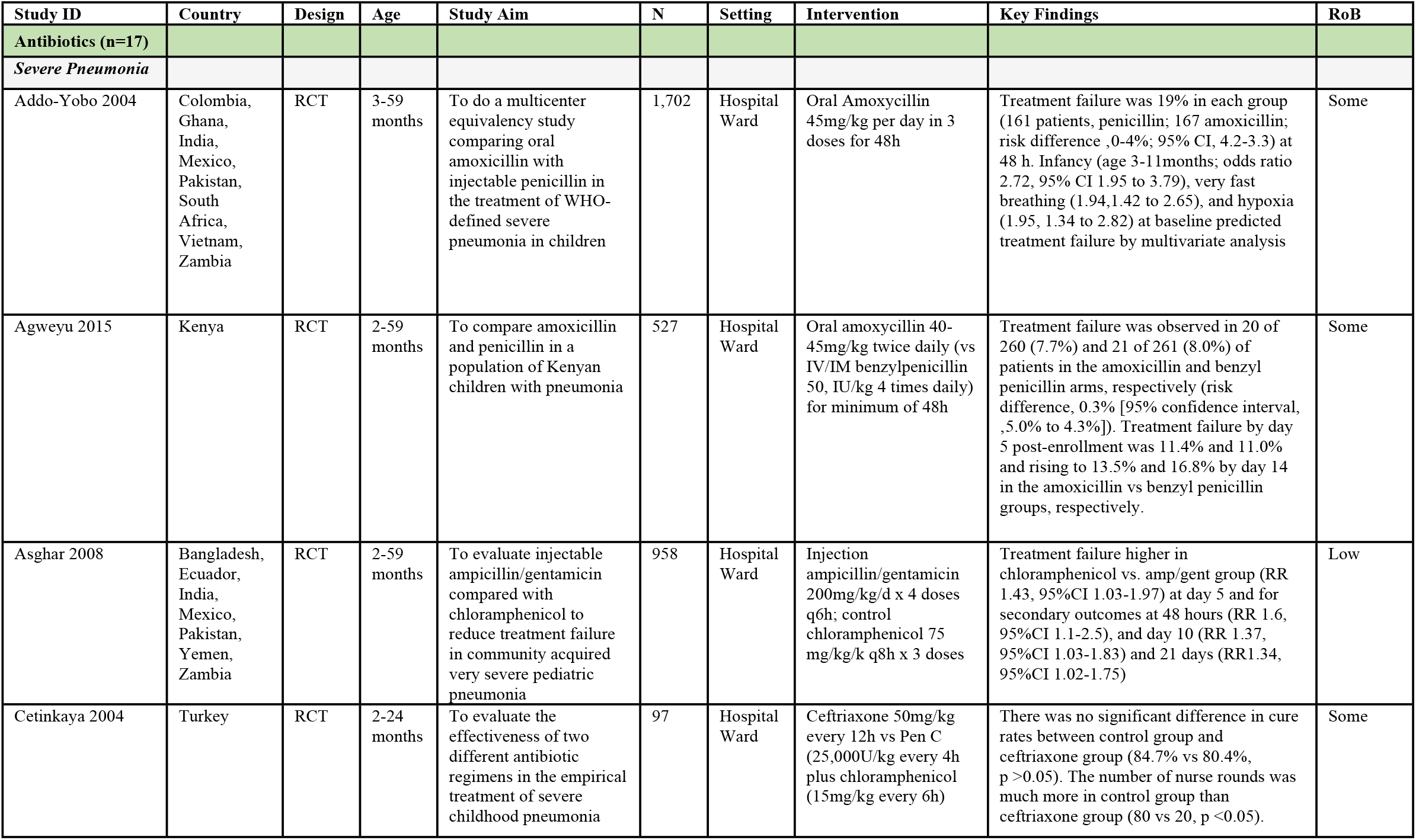
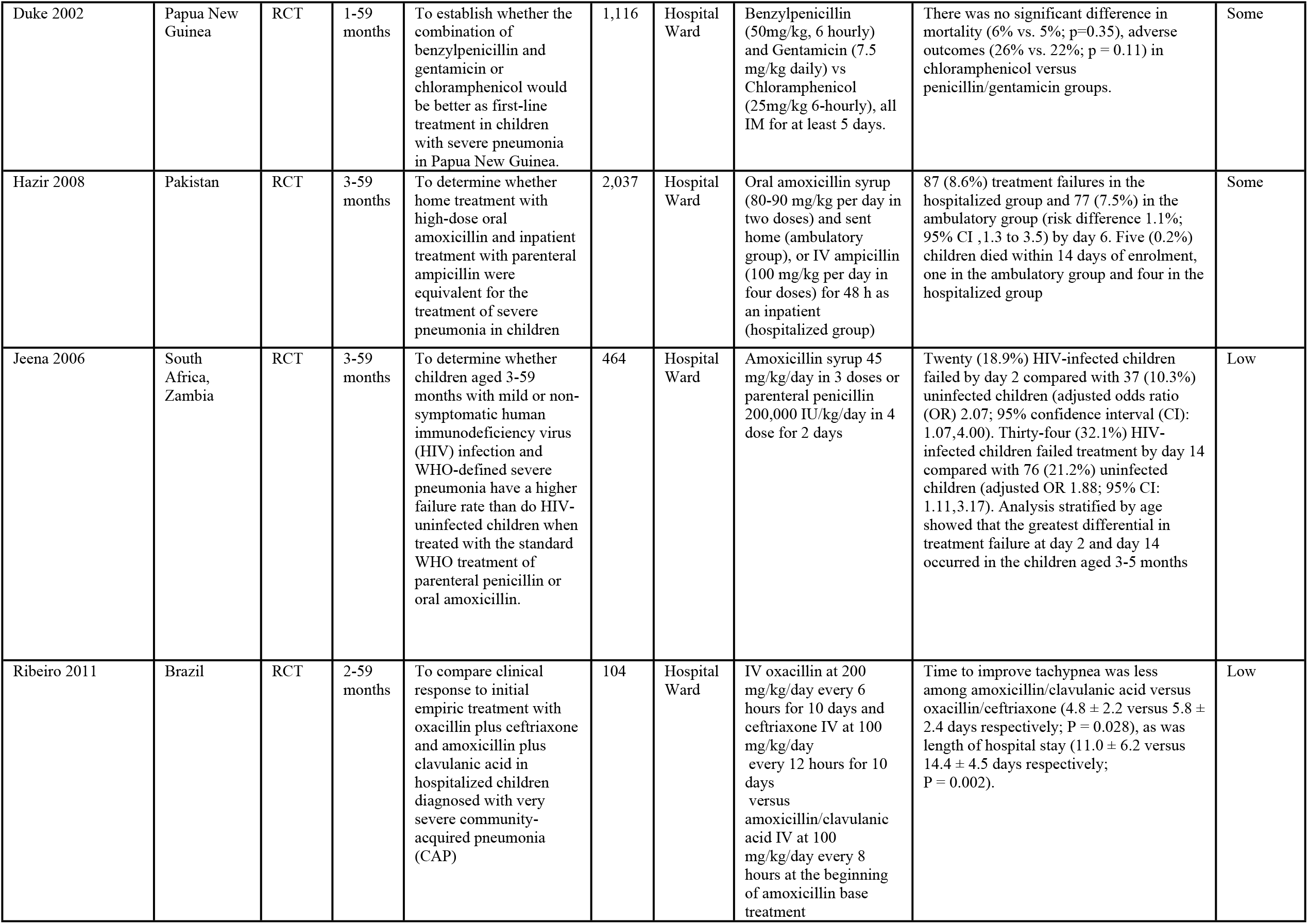
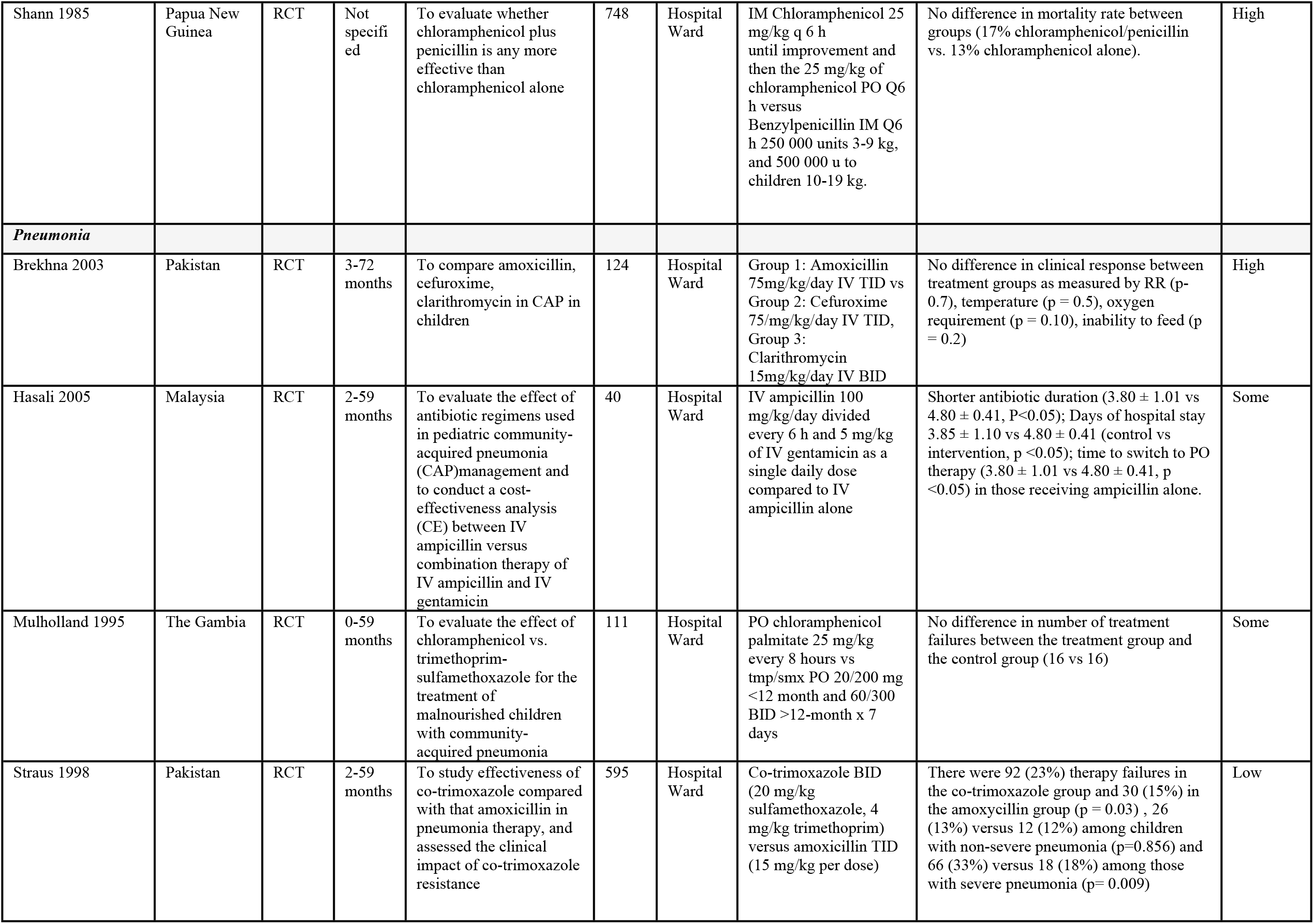
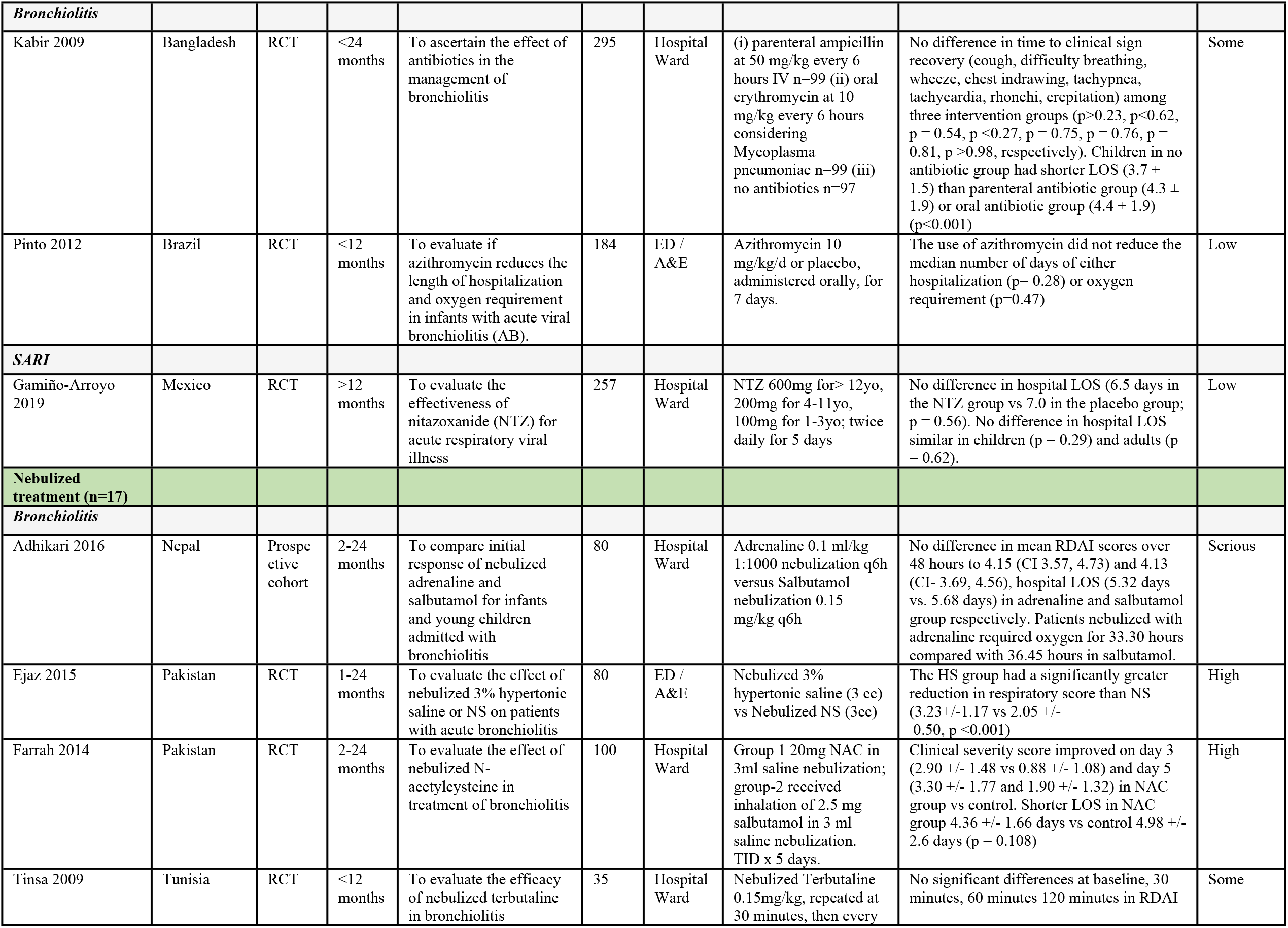
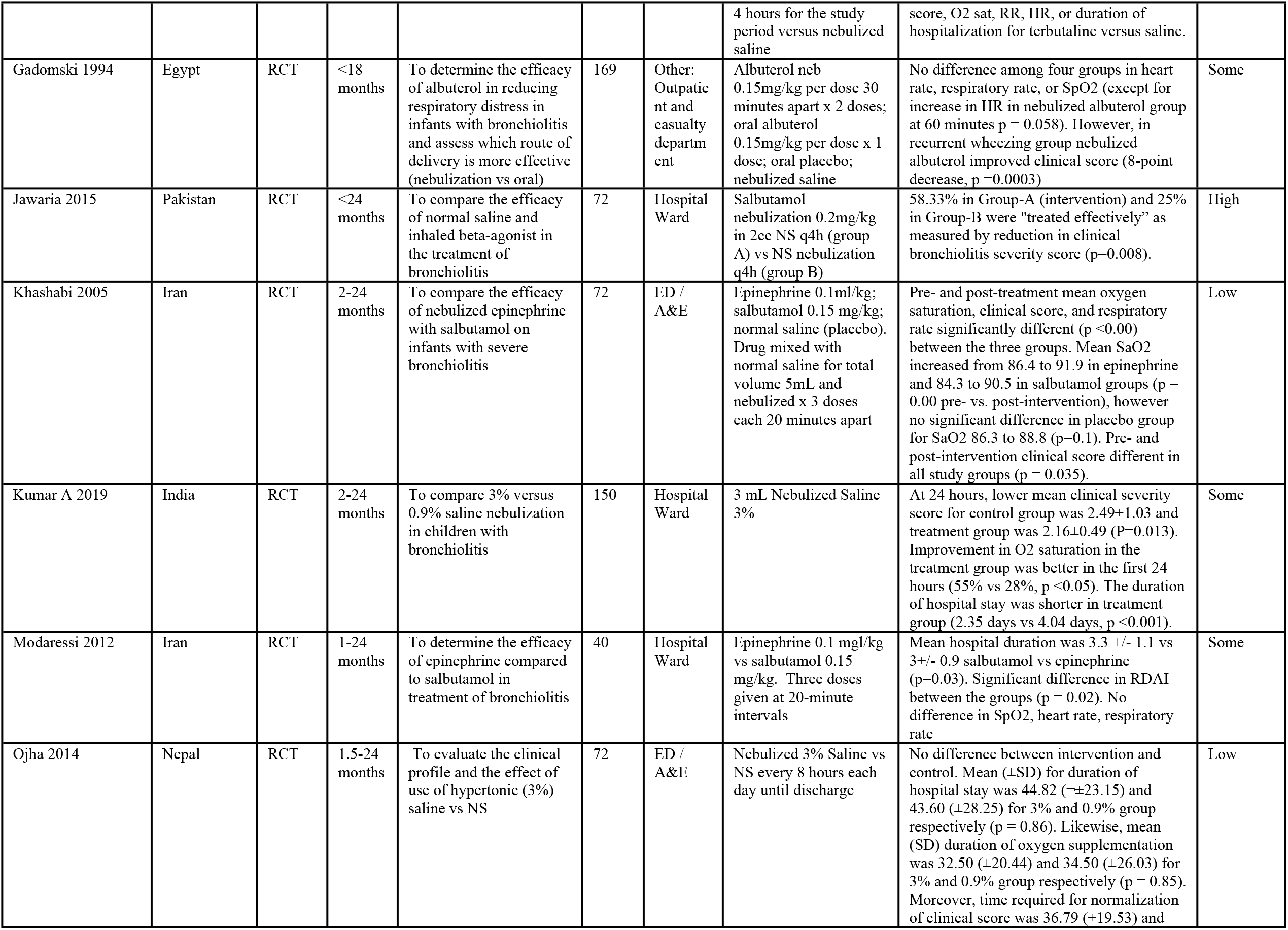
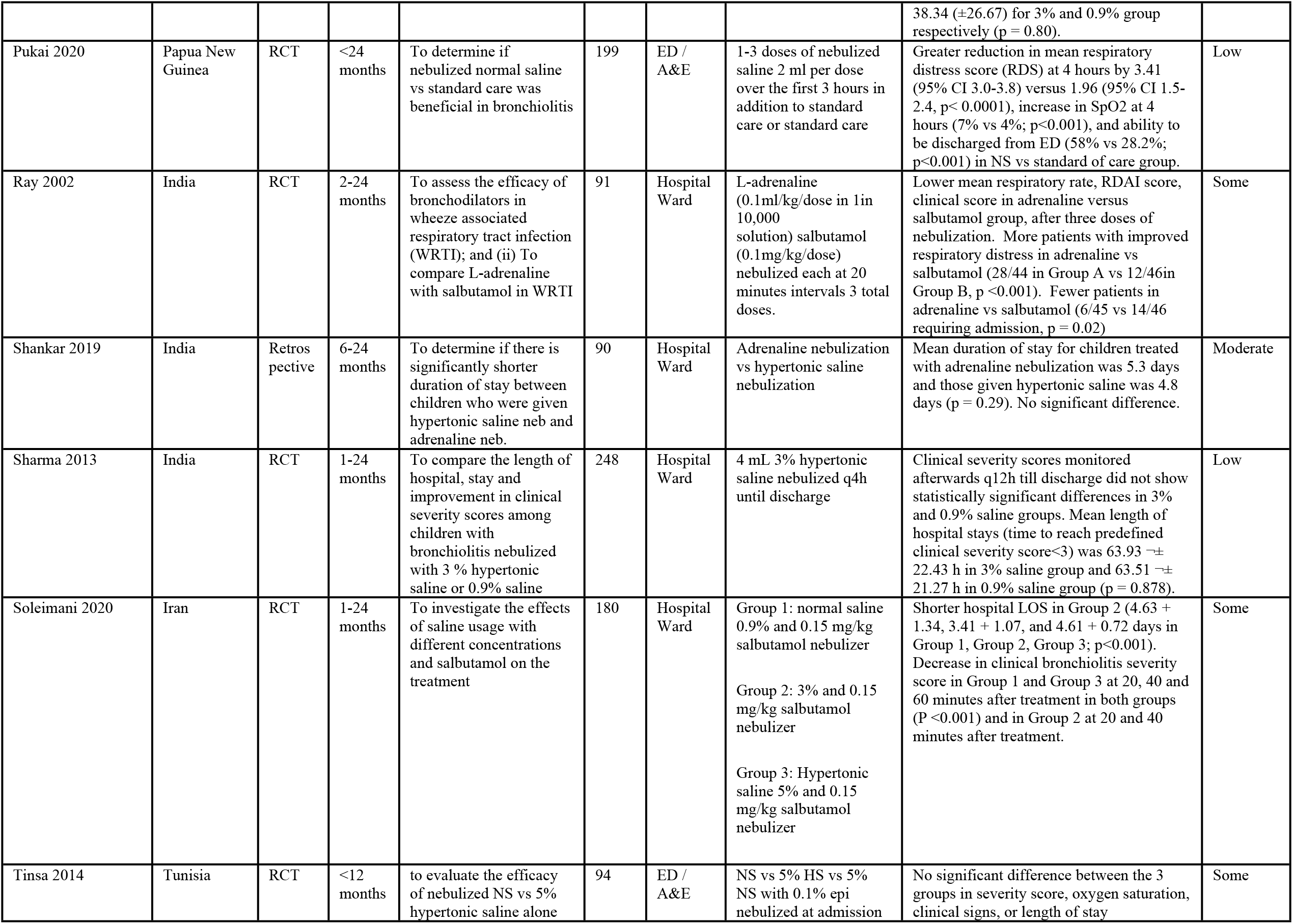
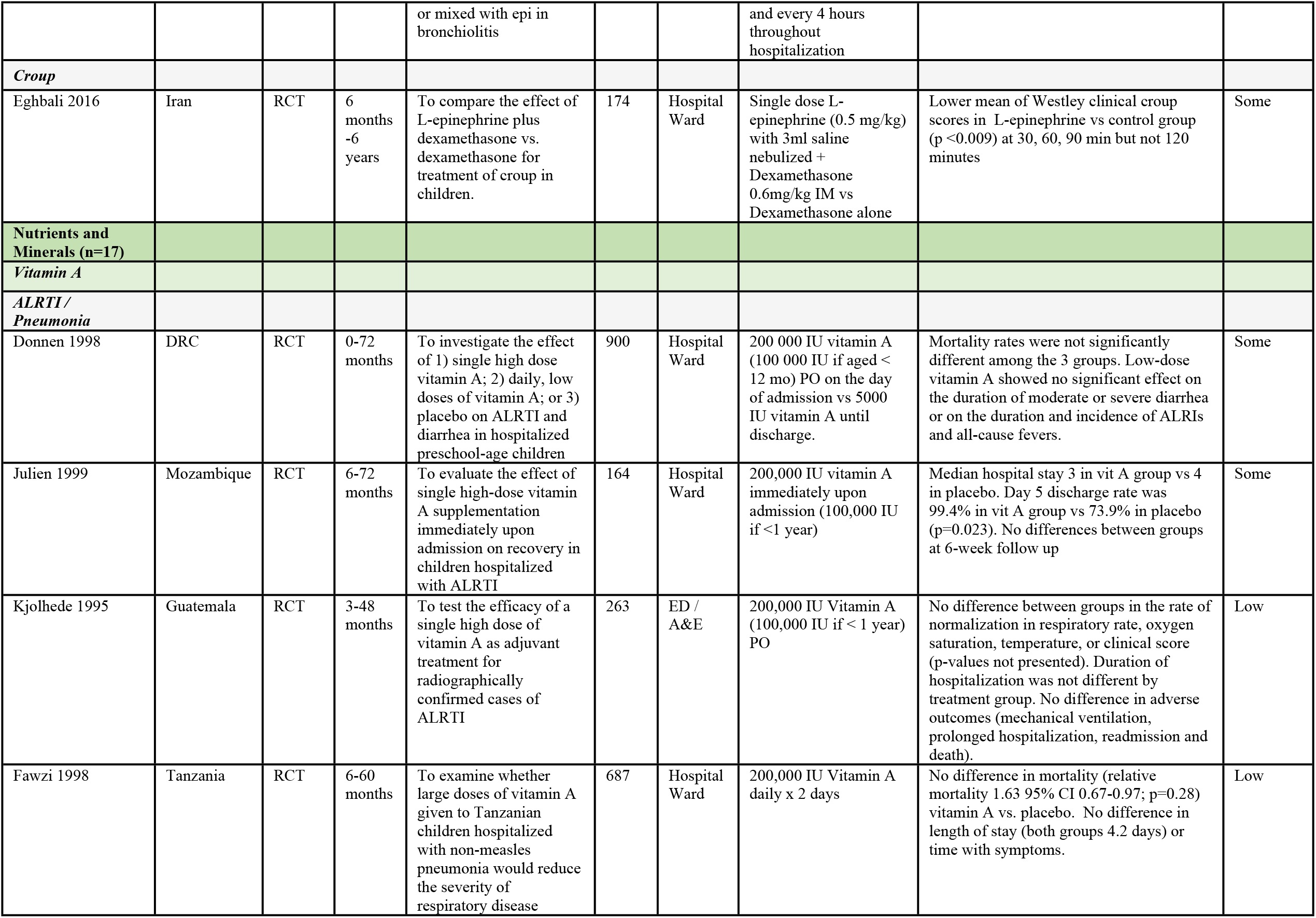
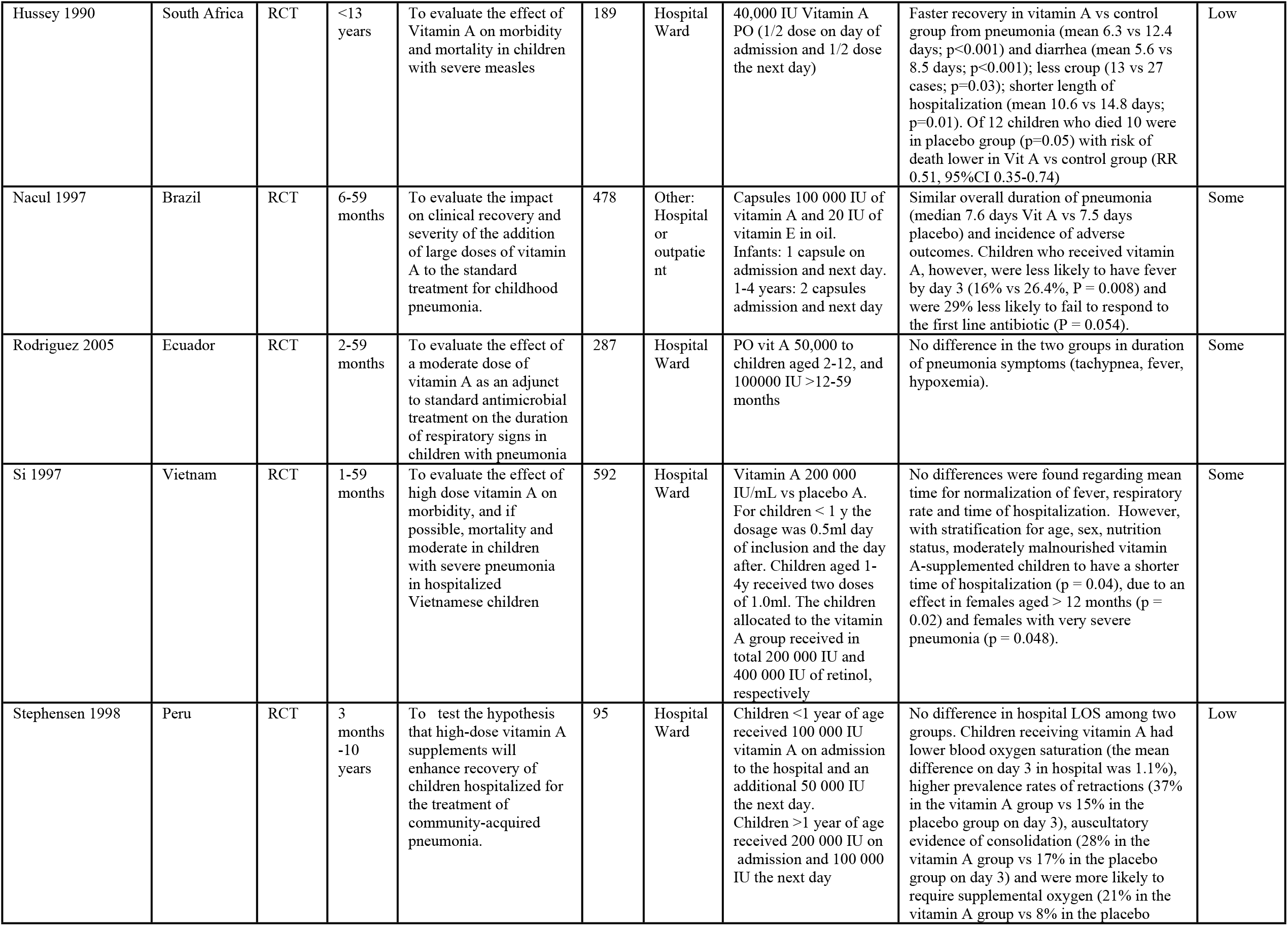
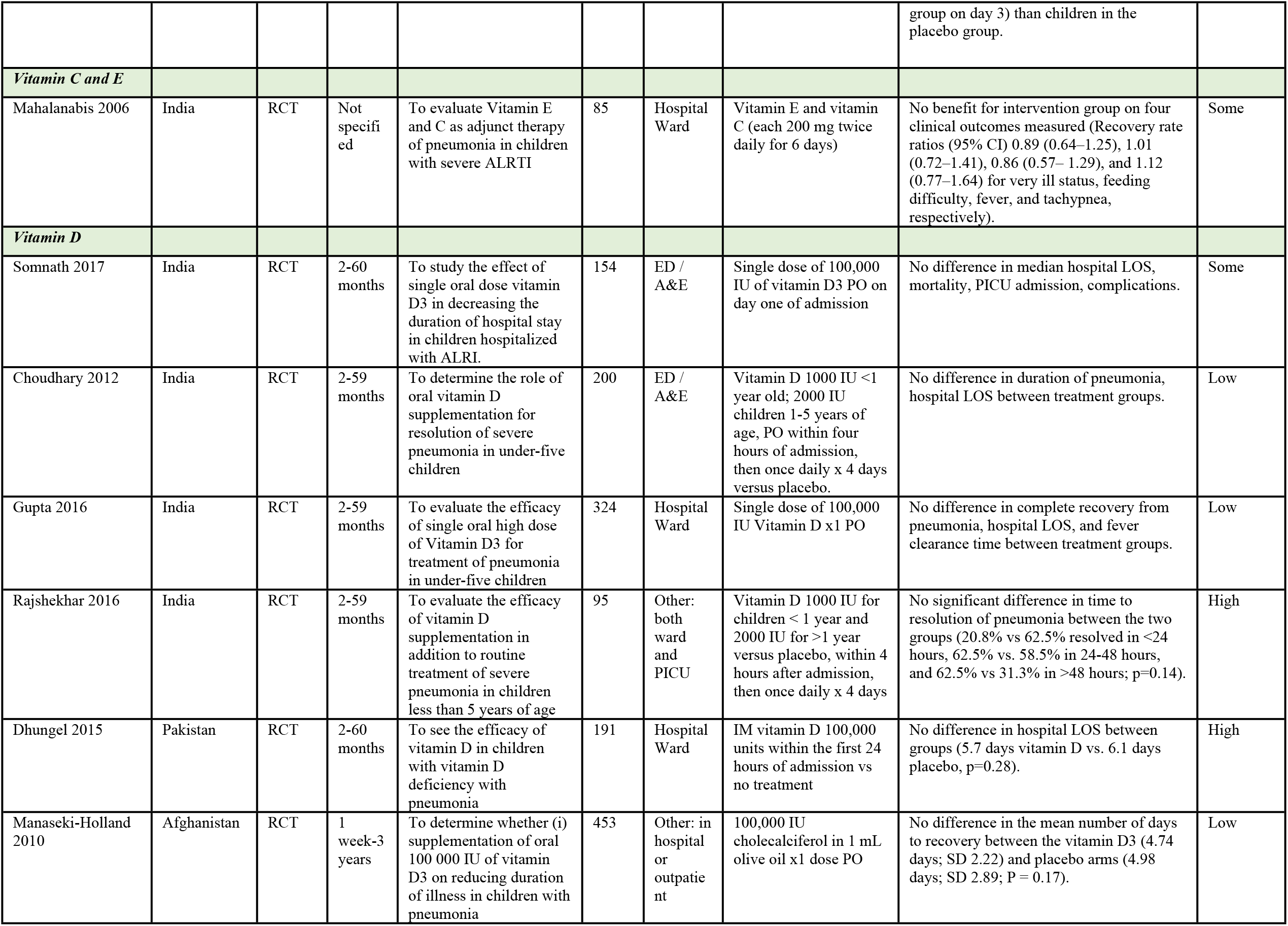
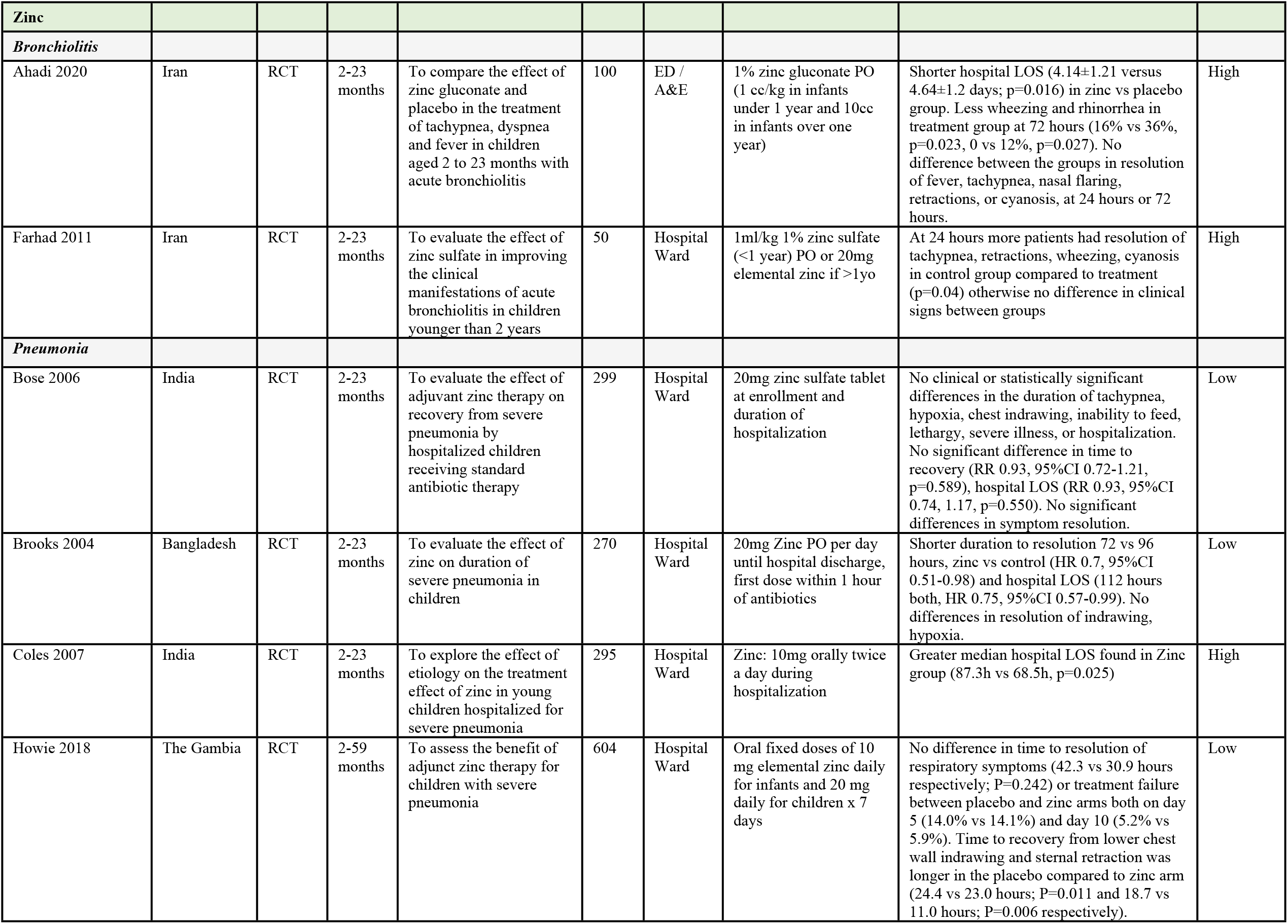
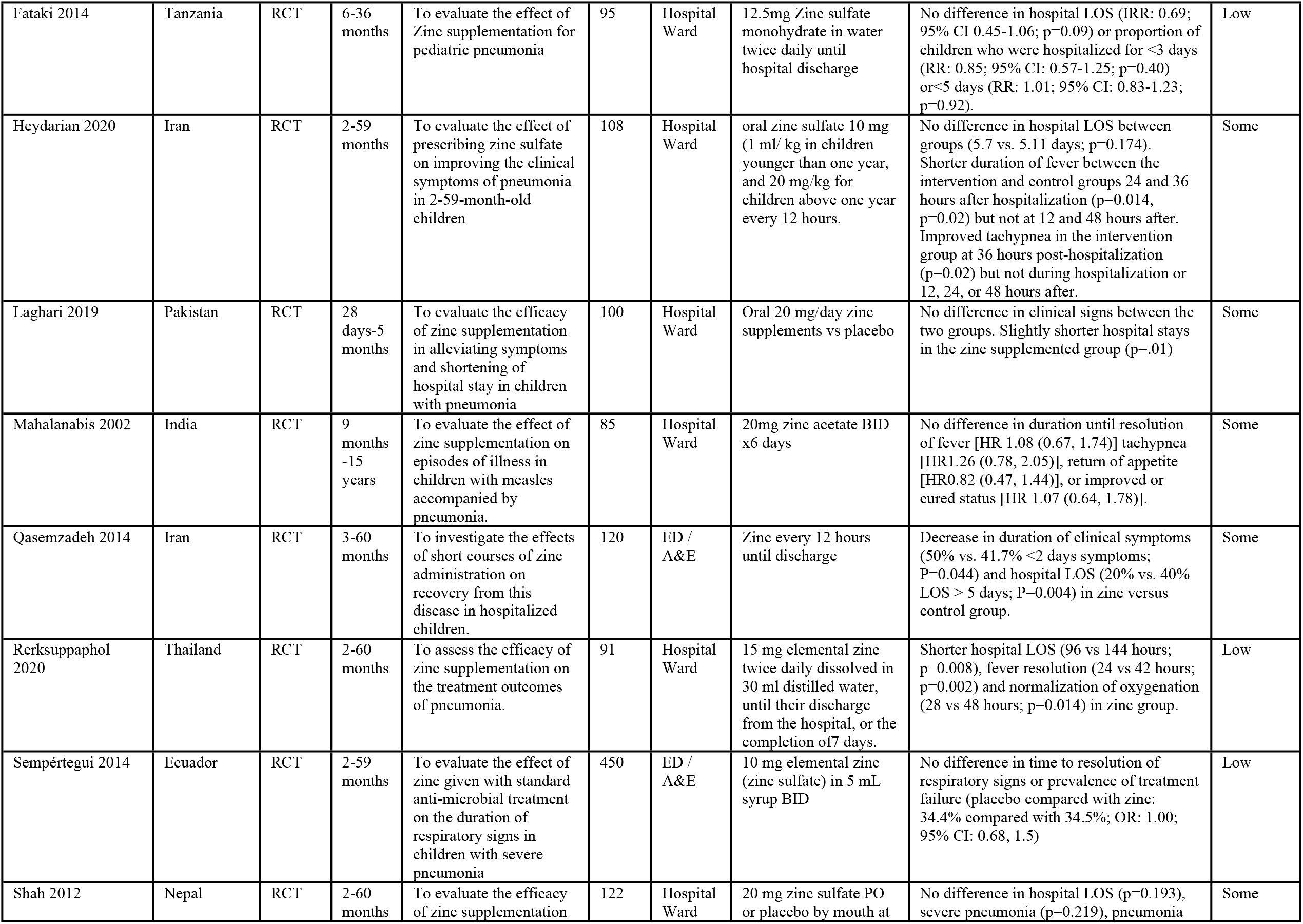
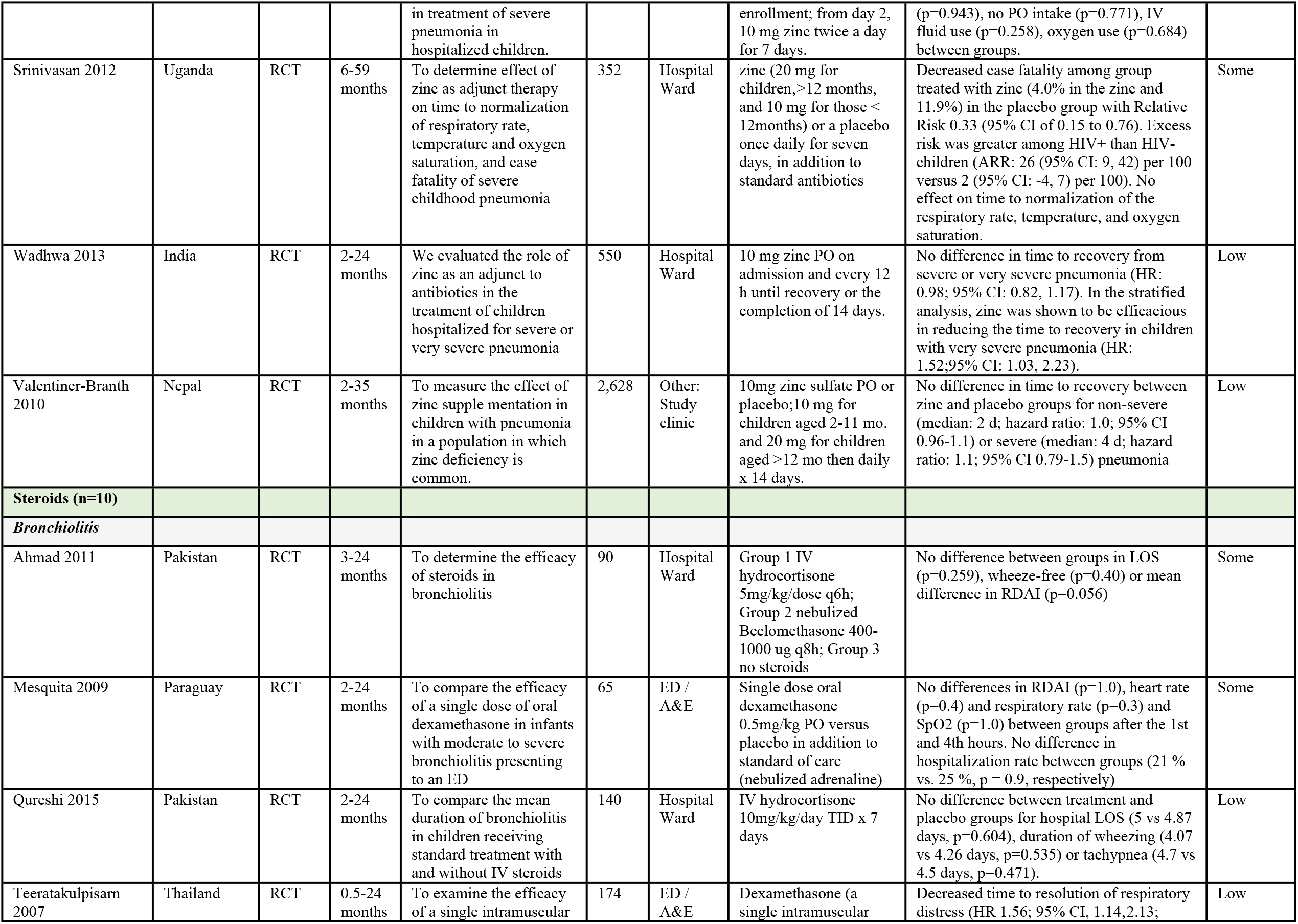
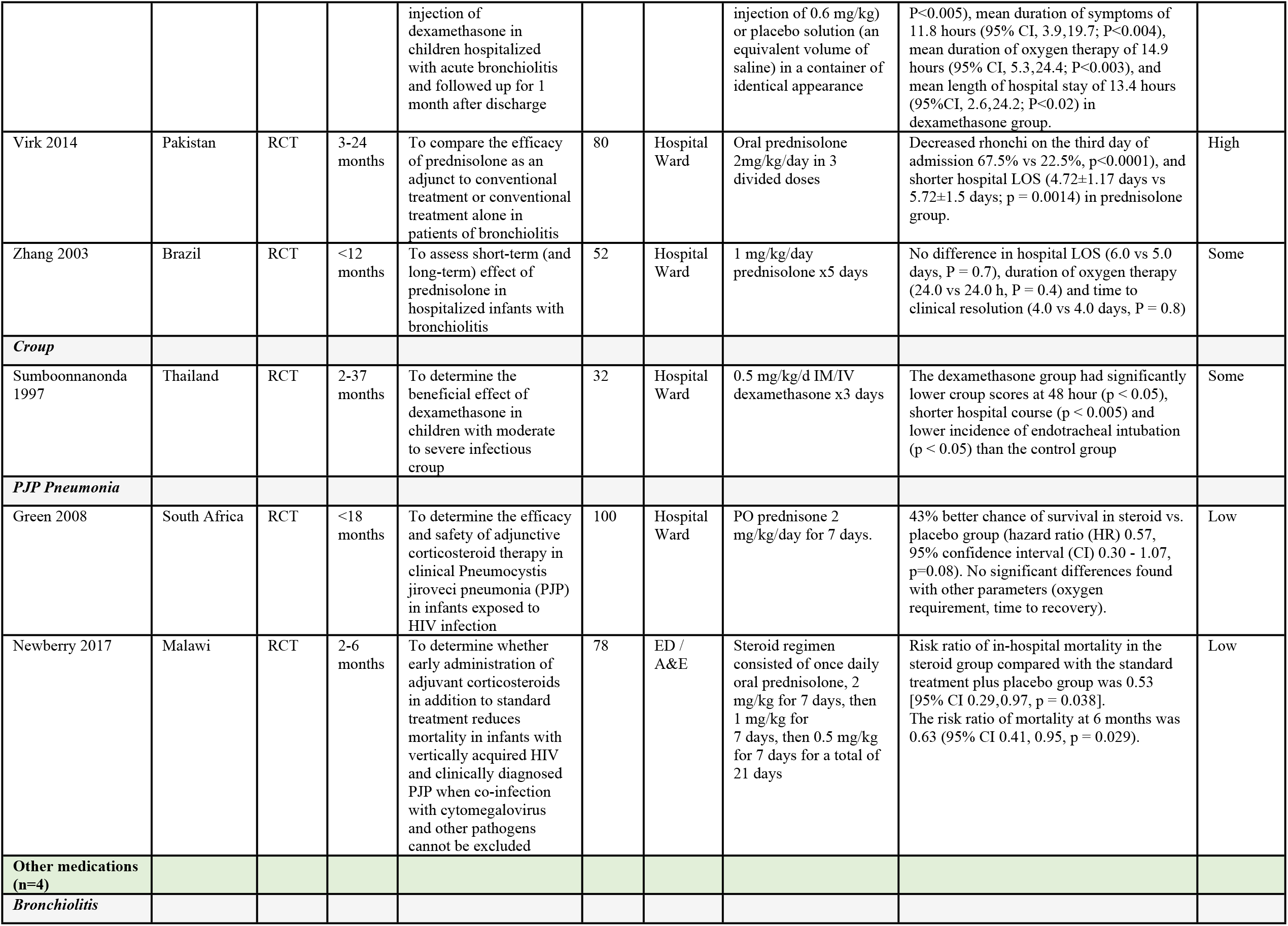
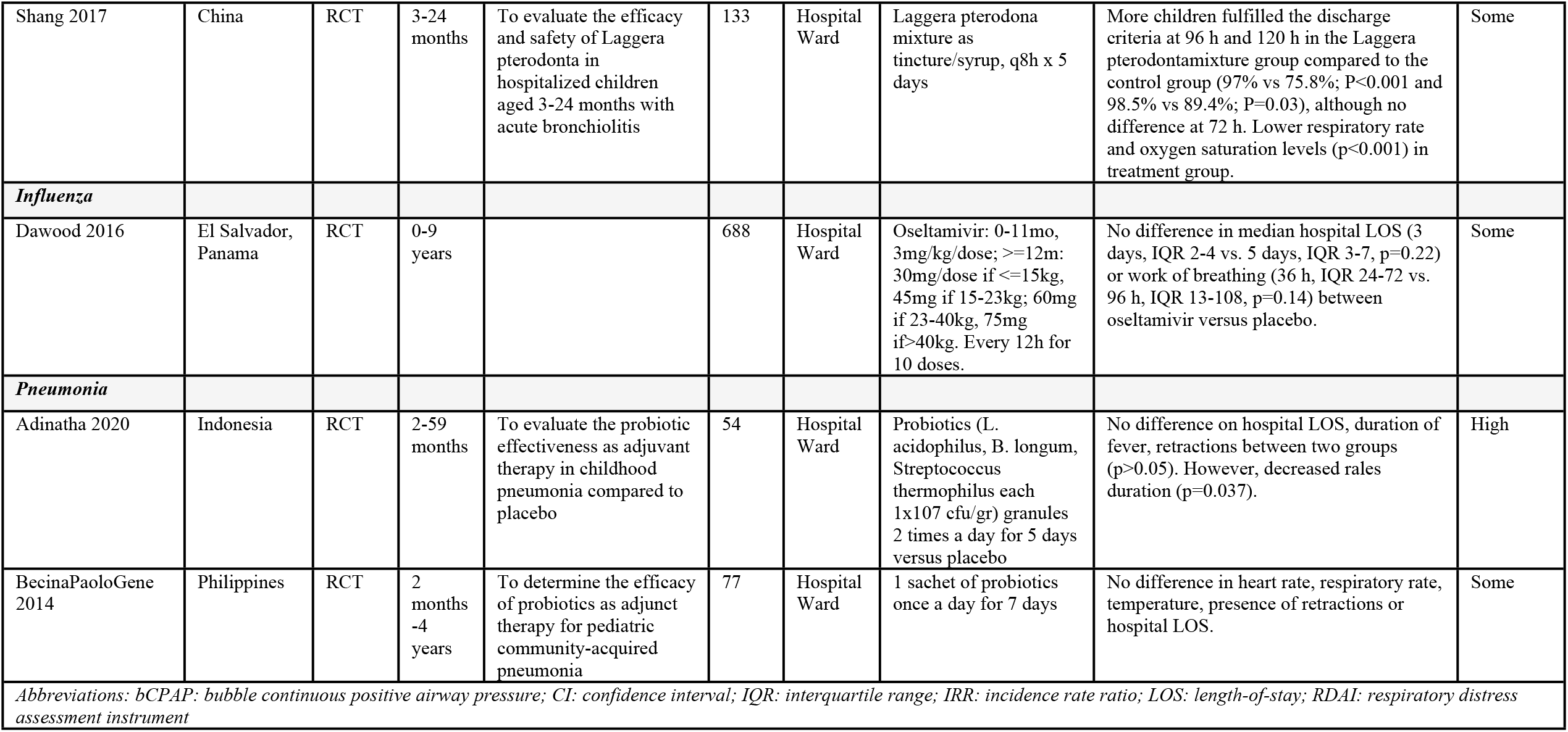
Summary of Findings – Medications (N=80).

**Figure 1.**
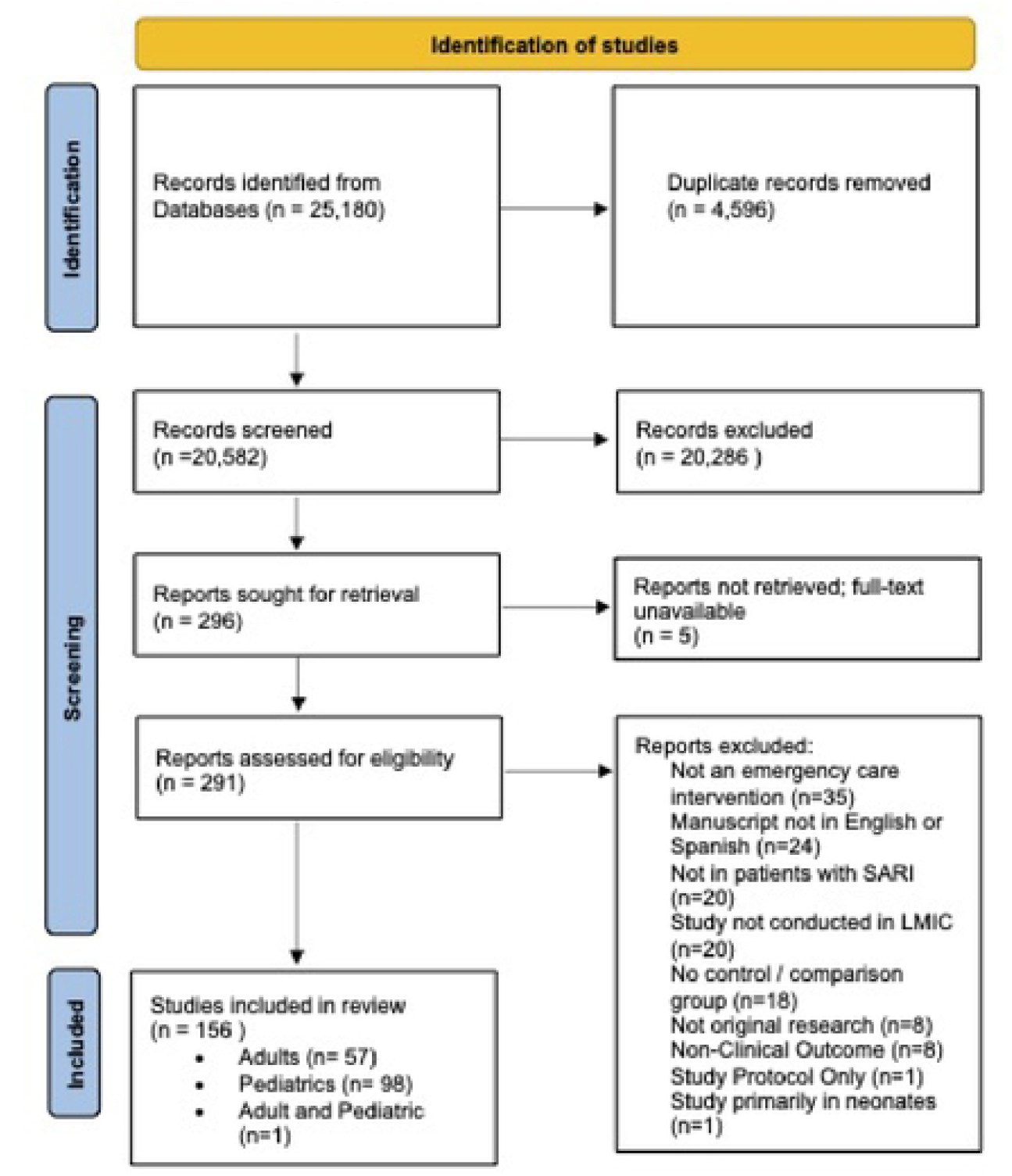
PRISMA flow diagram for systematic review.

**Figure 2.**
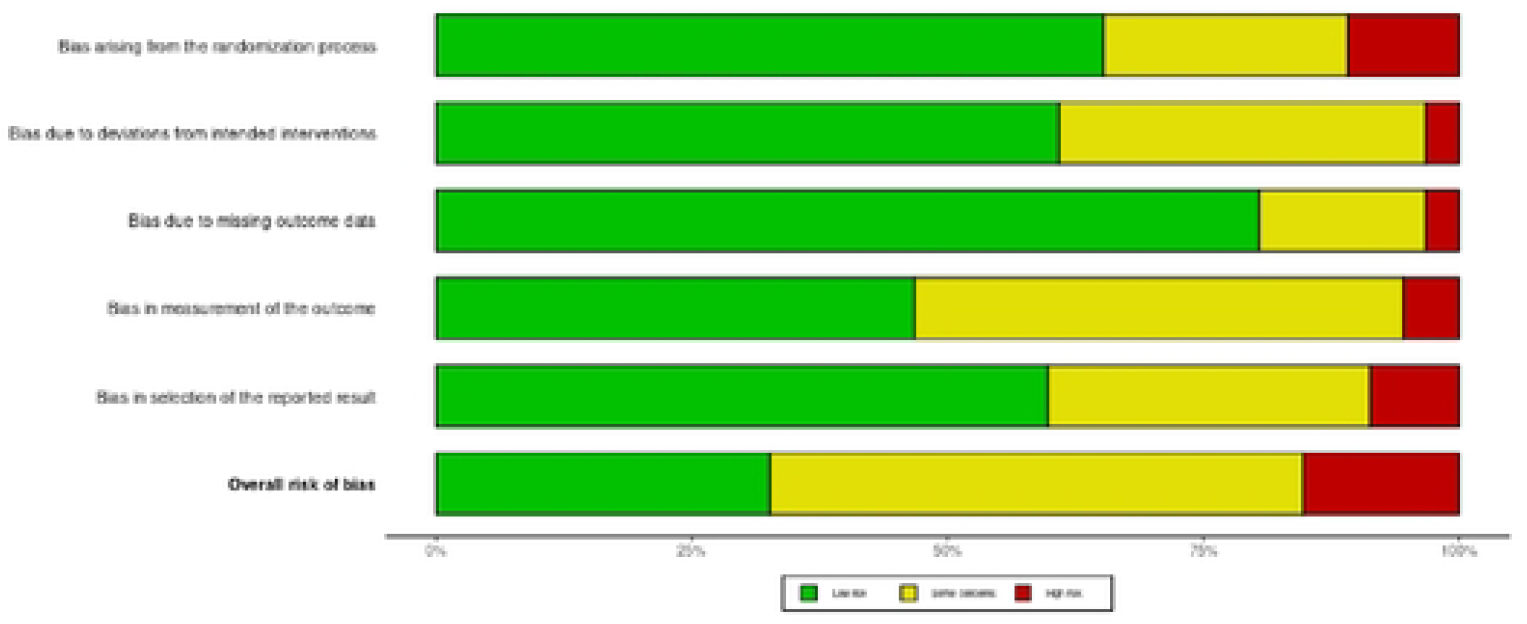
Summary of risk of bias assessments for included randomized-controlled trials using Cochrane risk-of-bias tool for randomized trials version 2 (RoB 2).

**Figure 3.**
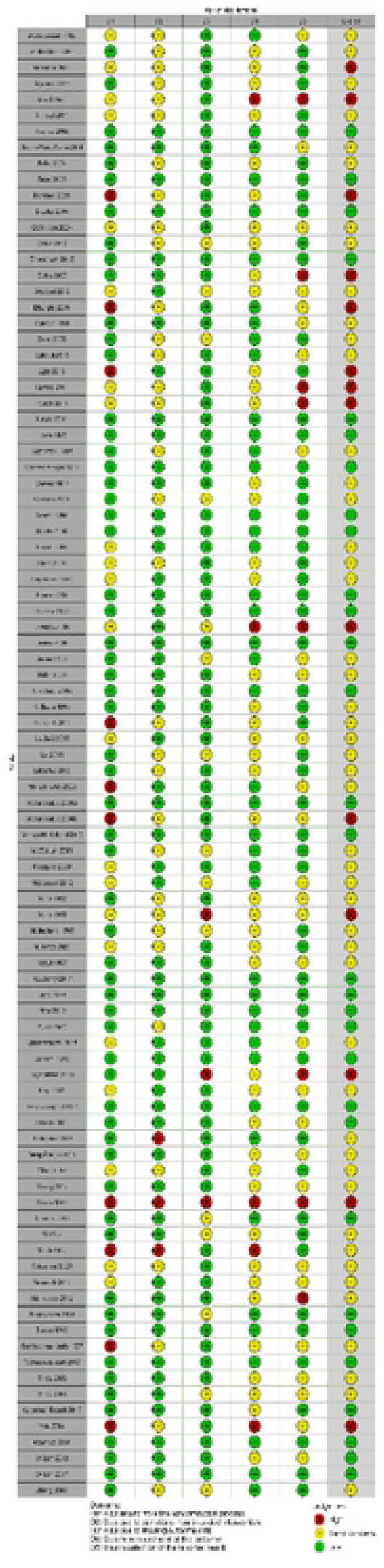
Consensus judgements on risk of bias for each included study. Each risk of bias item is assigned a color-coded ranking; green color represents low risk of bias, yellow some concerns, and red high risk of bias.

**Figure 4.**
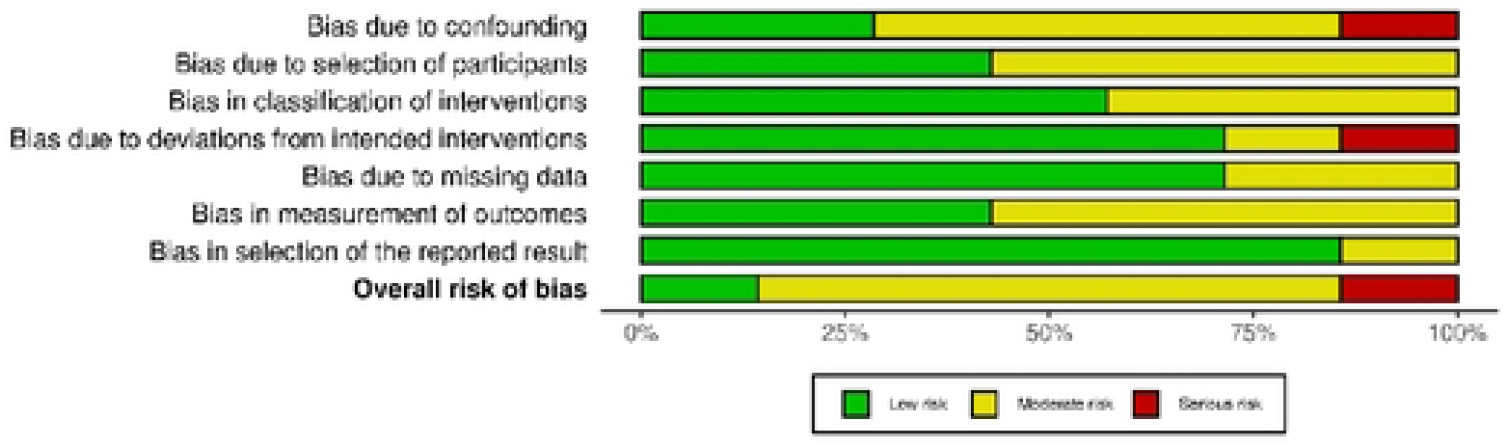
Summary of risk of bias assessments for included non-randomized studies according to the ROBINS-I tool.

**Figure 5.**
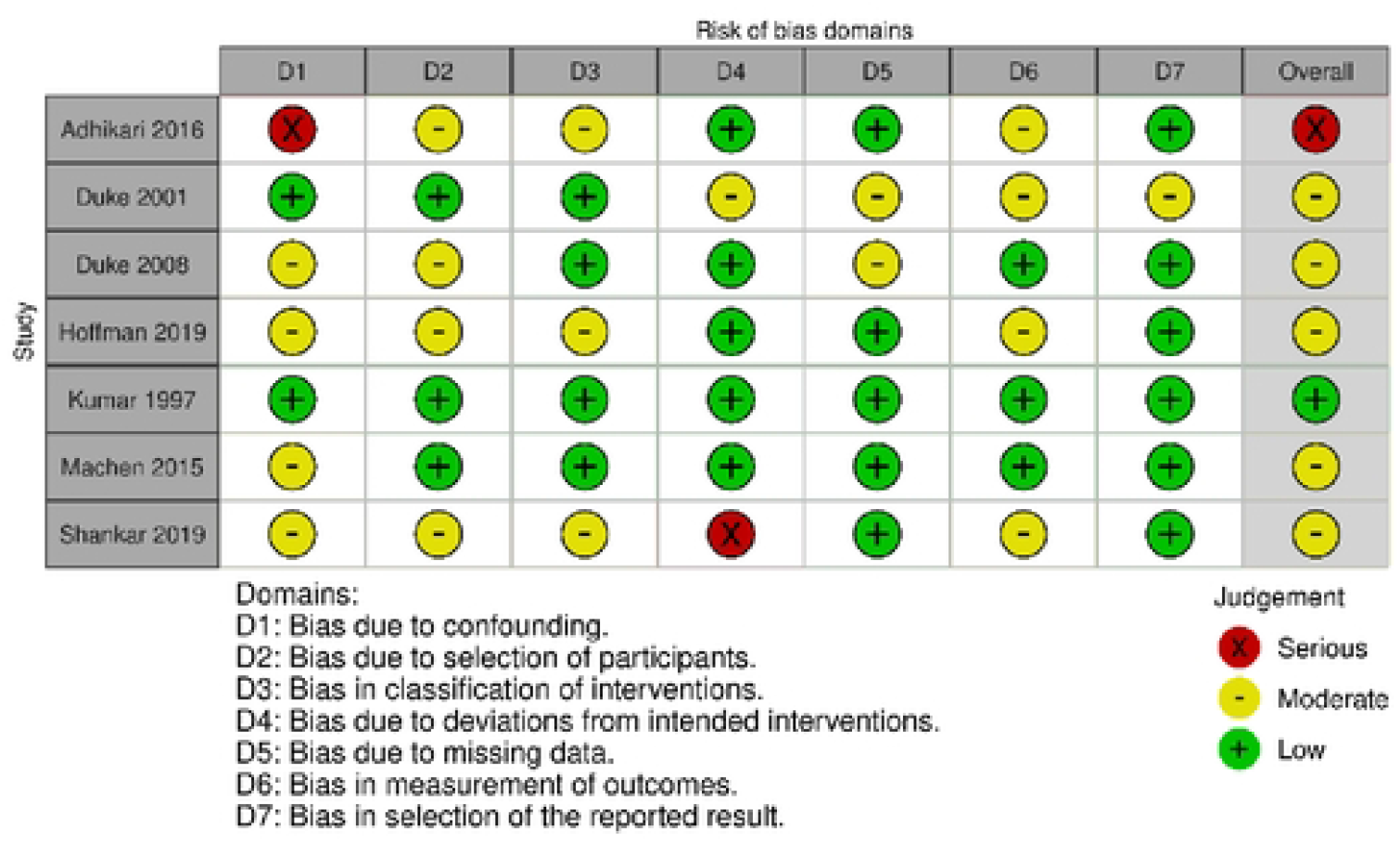
Consensus judgements using Cochrane ROBINS-I for each included nonrandomized study. Each risk of bias item is assigned a color-coded ranking; green color represents low risk of bias, yellow some concerns, and red high risk of bias.

### - AndRespiratory Support

Fourteen studies evaluated respiratory support interventions, including eight focused on oxygen delivery interventions and six on continuous positive airway pressure (CPAP).

#### Oxygen delivery, excluding CPAP

Eight studies evaluated various oxygen delivery systems, six on pneumonia or ALRTI and two on undifferentiated acute respiratory distress. Duke et al. (2001) conducted a prospective cohort study in Papua New Guinea which found a decreased mortality (but not statistically significant) among children monitored with pulse oximetry and administered oxygen if SpO2 < 85%. In another study by Duke et al. (2008), implementation of an oxygen administration guideline and oxygen concentrators was significantly associated with 35% lower risk of death. Mulondo et al. (Uganda) found that children with pneumonia treated with an oxygen-sparing nasal reservoir cannula had higher mean SpO2 without adverse events, although other clinical outcomes were not measured in this small pilot RCT. Three RCTs among children with ALRTI were included; Muhe et al. (Ethiopia) conducted two RCTs (in 1997 and 1998) on oxygen administration via nasal catheter versus nasal prongs and found no differences in hypoxia or complications in children with ALRTI, although authors recommended the use of nasal prongs as needed and nose bleeding/ulceration was greater for nasal catheter use. In a stepped-wedge cluster randomized trial, Graham et al. (Nigeria) found decreased odds of death among children with ALRTI treated in hospitals after implementation of multifaceted standardized oxygen system package (improved oxygen equipment, clinical education, technical training/support, infrastructure support) over pulse oximetry introduction or baseline (usual care prior to introducing pulse oximetry), although benefit was not seen in children overall.

Two studies evaluated interventions for undifferentiated respiratory distress; in a pre-post study, Hoffman et al. (South Africa) found that children treated during a period where high-flow nasal cannula was available may reduce need for transfers to higher level health facilities, although the study was retrospective and underpowered for mortality benefit. Kumar et al. (India) evaluated several modes of oxygen delivery and found improved treatment of hypoxia among children administered oxygen via a headbox compared to face mask or nasal catheter.

#### Continuous Positive Airway Pressure (CPAP), including Bubble CPAP

Two RCTs evaluated bubble CPAP (bCPAP) for pneumonia. Chisti et al. found that among children with pneumonia in Bangladesh, those who received bCPAP had lower mortality rate (4% vs. 15%; RR 0.25; 95% CI 0.07-0.89) and lower treatment failure compared to low-flow oxygen therapy. However, McCollum et al. (Malawi) found that bCPAP was associated with higher hospital mortality (17% vs 11%; RR 1.52, 95% CI 1.02-2.27) compared with low flow oxygen in children, suggesting that bCPAP might carry risks not previously recognized. Three studies evaluated bCPAP for undifferentiated respiratory distress; an RCT by Wilson et al. (2013) in Ghana found decreased respiratory rate by 16 breaths/minute in children treated with immediate initiation (<1 hour) of CPAP, although no change in children who had CPAP delayed over an hour. Another RCT by Wilson et al. (2017) found no difference in mortality at two weeks in unadjusted analysis, however found a benefit with decreased mortality among children less than one year of age. Machen et al. (Malawi) evaluated bCPAP in a non-randomized study among children with undifferentiated acute respiratory distress in Malawi and found the best outcomes among children with bronchiolitis (92.9% survival), followed by pneumonia (53.3% survival), and PJP pneumonia (38.1% survival) although the study was limited by a lack of randomization and comparison of clinical outcomes. Lal et al. (India) found improved respiratory rate and clinical respiratory distress scores (Silverman- Anderson and Modified Pediatric Society of New Zealand scores) at one hour in children with bronchiolitis treated with nasal CPAP compared to oxygen through mask or hood in India.

### Supportive Care

Five studies evaluated supportive care interventions, including four on chest physiotherapy and one on steam therapy.

#### Physiotherapy

Four RCTs evaluated the effectiveness of chest physiotherapy (PT); two for pneumonia and two for bronchiolitis^9-12^. Lukrafka et al. (Brazil) and Bohe et al. (Argentina) found no differences in severity score nor hospital LOS among children with pneumonia treated with chest PT. However, Abdelbasset et al. (Egypt) found faster time to clinical resolution of pneumonia and improvement in respiratory rate in children in the chest PT group and Gomes et al. (Brazil) found improvement in Wang respiratory score in infants with bronchiolitis treated with “new” chest physical therapy (with prolonged slow expiration and clearance rhinophayngeal retrograde) or conventional chest PT compared with suction of upper airways alone.

#### Other supportive care interventions

Singh et al. found that steam therapy had no benefit in children with pneumonia compared to standard of care although improvement was seen in children with bronchiolitis^13^.

### Medications – Antibiotics

Seventeen RCTs evaluated antibiotic-based interventions; nine on severe pneumonia, five on pneumonia, two RCTs on bronchiolitis and one on SARI generally.

#### Antibiotics for Severe Pneumonia

Four RCTs evaluated oral amoxicillin versus parenteral antibiotics for severe pneumonia^14-17^. Three studies found oral amoxicillin versus injectable penicillin or ampicillin to have equal efficacy for children 3-59 months of age^14,15,17^. In a sub-analysis of the trial conducted by Addo Yobo et al, Jeena et al. found greater treatment failure with oral amoxicillin or parenteral penicillin at day 2 and day 14 among children with Human Immunodeficiency Virus (HIV) compared with children without HIV.

Five RCTs evaluated various parenteral antibiotic regimens, four of which included chloramphenicol. Asghar et al. found greater treatment failure with chloramphenicol compared to ampicillin/gentamicin in children aged 2-59 months across seven LMICs. No difference in cure rates was found with ceftriaxone versus penicillin plus chloramphenicol (Cetinkaya et al.), mortality rates with benzylpenicillin plus chloramphenicol versus chloramphenicol alone (Shann et al.), or mortality rates and adverse outcomes in penicillin/gentamicin versus chloramphenicol (Duke et al.). Ribeiro et al. (Brazil) found hospital LOS and time to improvement of tachypnea was shorter with oxacillin/ceftriaxone versus amoxicillin/clavulanic acid in hospitalized children diagnosed with very severe pneumonia.

#### Antibiotics for Non-Severe Pneumonia

Four RCTs evaluate various antibiotic regimens for non-severe pneumonia. Straus et al. (Pakistan) found more treatment failures with trimethoprim-sulfamethoxazole (TMP-SMX) versus amoxicillin. Mulholland et al. (The Gambia) found no difference in treatment failure with chloramphenicol versus TMP-SMX for the treatment of malnourished children with community-acquired pneumonia. Hasali et al. (Malaysia) found shorter hospital LOS and time to switching to PO antibiotics with ampicillin-gentamicin combination compared to ampicillin alone. Brekhna et al. (Pakistan) found no difference in clinical response as defined as oxygen requirement, feeding ability, or tachypnea treated with amoxicillin versus cefuroxime versus clarithromycin, although this study was deemed to have a high risk of bias.

#### Antibiotics for Bronchiolitis

Kabir et al. (Bangladesh) found no benefit for parenteral ampicillin versus oral erythromycin versus no antibiotics on recovery of clinical symptoms in children with bronchiolitis; furthermore, children in the group given no antibiotics had a shorter hospital LOS compared with those who received antibiotics (3.7 days vs 4.4 days). Pinto et al. (Brazil) found that azithromycin had no effect on the duration of hospitalization or oxygen requirement in children with bronchiolitis under one year.

#### Antibiotics for Undifferentiated SARI

Gamiño-Arroyo et al. evaluated the effectiveness of nitazoxanide for SARI in patients over 12 months of age in Mexico and found no difference in hospital LOS and no difference in viral shedding.

### Medications – Nebulized Treatments

Seventeen studies evaluated nebulized treatments including 16 on bronchiolitis and one on croup.

#### Nebulized Treatments for Bronchiolitis

Six studies evaluated different concentrations of nebulized saline for children with bronchiolitis. Five RCTs evaluated hypertonic (3%) saline versus normal (0.9%) saline (NS); three studies found no benefit for 3% saline on clinical status or hospital LOS^18-20^. Two RCTs found benefit for 3% saline. However, Ejaz et al. (Pakistan) found greater reduction in respiratory score in the 3% saline group while Kumar et al. (India) found greater improvement in clinical severity, oxygen saturation at 24 hours, and shorter hospital length of stay with 3% saline. Soleimani et al. (Iran) compared three different concentrations of nebulized saline combined with salbutamol and found that in children 1-24 months those treated with 3% saline had shorter hospital LOS compared to those treated with 5% or NS.

Two studies evaluated nebulized salbutamol versus epinephrine with mixed results; Ray et al. (India) found lower clinical severity score and respiratory rate for salbutamol over epinephrine, while Adhikari et al. (Nepal) found no difference in respiratory distress assessment instrument (RDAI) scores between those treated with epinephrine versus salbutamol. Three studies evaluated bronchodilators versus nebulized saline, two of which found benefit for bronchodilators; Jawaria et al. (Pakistan) found that more children treated with salbutamol had a reduction in clinical bronchiolitis severity score compared to NS, although the study had high risk of bias; Khashabi et al. (Iran) found improved oxygen saturation, clinical score and respiratory rate in children treated with nebulized salbutamol or epinephrine compared to NS; however, Tinsa et al. (2009; Tunisia) found no difference in RDAI score or hospital LOS for nebulized terbutaline over saline. Shankar et al. (India) found no significant difference in clinical severity scores or hospital LOS for nebulized hypertonic saline versus epinephrine in India. Gadomski et al. (Egypt) compared nebulized albuterol, nebulized saline, orally administered albuterol, and orally administered placebo in Egypt and found no difference in outcomes among the four groups, except for an improvement in respiratory rate among those with history of recurrent wheezing. Farrah et al. (Pakistan) found no difference in clinical severity score with nebulized NAC versus salbutamol. Pukai et al. (Papua New Guinea) found that children less than 24 months with either bronchiolitis or pneumonia treated with NS versus standard of care had a greater reduction in respiratory distress score (RDS), and increase in pulse oximetry at 4 hours, and ability to be discharged from the ED.

#### Nebulized Treatments for Croup

Eghbali et al. (Iran) found a reduction in Westley clinical croup scores for nebulized epinephrine plus dexamethasone vs. nebulized saline plus dexamethasone in children aged six months to six years at 30, 60, and 90 minutes, although this difference was not seen at 120 minutes.

### Medications – Nutrients and Minerals

Seventeen studies evaluated nutrients and minerals (Vitamin A, C / E, D, and Zinc) as an intervention for treatment of children with acute lower respiratory tract infections (ALRTI), including pneumonia and bronchiolitis.

#### Vitamin A for Acute Lower Respiratory Tract Infections (ALRTI), including Pneumonia

Three RCTs evaluated a single high dose of vitamin A among hospitalized children with ALRTIs. Donnen et al. (Democratic Republic of the Congo) found no effect on mortality rates of a high-dose vitamin A versus low dose daily vitamin A on children with ALRTIs or diarrhea. Similarly, Kiolhede et al. (Guatemala) found no effect of adjuvant high dose vitamin A versus placebo on clinical parameters (e.g., respiratory rate, oxygen saturation), hospital LOS, or death. However, Julien et al. (Mozambique) found a shorter median hospital LOS (4 days vs. 3 days) among children treated with single high-dose vitamin A.

Six RCTs evaluated vitamin A for children with pneumonia^21-26^. Four RCTs evaluated high-dose vitamin A given on the day of admission and an additional dose on the subsequent day, three of which (conducted in Tanzania, Peru, and Brazil) found no effect on mortality, hospital LOS, or duration of pneumonia symptoms^22,24,26^. Furthermore, Stephensen et al. (Peru) additionally found greater adverse effects including lower oxygen saturation in children treated with high-dose vitamin A with pneumonia in Peru. However, Si et al. (Vietnam) found while there was no effect on hospital LOS in children with moderate-to-severe pneumonia overall, although a shorter hospital LOS among malnourished children (particularly females with very severe pneumonia) treated with high-dose vitamin A. Rodriguez et al. found no difference with treatment with moderate-dose vitamin A on duration of pneumonia in children with non-measles pneumonia in Ecuador. Lastly, Hussey et al. Focused on patients specifically with measles pneumonia and found lower mortality, shorter hospital LOS, and faster recovery in children who received vitamin A in South Africa

#### Vitamin C and E for ALRTI, including pneumonia

Mahalanabis et al. (India) evaluated Vitamin C and E together as adjunctive treatment for pneumonia in India and found no significant benefits of the combination therapy compared to placebo on tachypnea, fever, feeding or clinical status^27^.

#### Vitamin D for ALRTI, including pneumonia

Six RCTs evaluated the effect of vitamin D in children under five years with ALRTI or pneumonia, all of which found no benefit for vitamin D^28-33^. Somnath et al. (India), Gupta et al. (India), Dhungel et al. (Pakistan), Choudhary et al. (India), and Manaseki-Holland et al (Afghanistan) found no effect of a single high dose of vitamin D on hospital LOS or mortality in children under five years hospitalized with ALRTI. Similarly, Rajshekhar et al. (India) found no difference in time to resolution of pneumonia symptoms.

#### Zinc for Bronchiolitis

Two RCTs evaluated the effect of zinc on bronchiolitis in children under two years^34,35^. Ahadi et al. found that children in the treatment group had shorter hospital LOS (4.14 ± 1.21 versus 4.64 ± 1.2 days; p=0.016) and less wheezing and rhinorrhea at 72 hours (16% vs 36%, p=0.023, 0 vs 12%, p=0.027, respectively). Both found no difference in tachypnea, retractions, or cyanosis at 24 hours in the treatment versus control group; both studies were deemed to have high risk of bias.

#### Zinc for Pneumonia

The effect of zinc on pneumonia outcomes were evaluated in 15 RCTs^36-50^. Seven studies found no benefit for zinc treatment over placebo^36,40,42,43,46,47,50^. Five studies reported favorable outcomes for zinc; Brooks et al. (Bangladesh) found a shorter duration to resolution of pneumonia and a one-day shorter hospital LOS in the zinc group (72 vs 96 hours; HR (0.7; 95% CI 0.51-0.98), but no differences in resolution of chest indrawing or hypoxia; Heydarin et al. (Iran) found no benefit for zinc on hospital LOS but found reported shorter duration of fever at 24 and 36 hours and improvement in tachypnea at 36 hours; Qasemzadeh et al. (Iran) and Rerksuppaphol et al. (Thailand) found shorter hospital LOS and time to resolution of clinical symptoms; Srinivasan et al. (Uganda) found decreased case fatality rate in children with severe pneumonia treated with zinc (4.0% vs. 11.9%), with greater effects in HIV-infected children. Conversely, Coles et al. (India) reported an increase in the median hospital LOS in the zinc group although the study had high risk of bias. Howie et al. (The Gambia) reported mixed outcomes with no difference found in time to resolution of symptoms, but marginal benefit for reduced time to resolution of chest indrawing and sternal retraction with zinc. Wadhwa et al. (India) found no benefit for zinc in their overall study population with severe pneumonia, however found benefit in stratified analysis among those with very severe pneumonia.

### Medications – Steroids

Nine studies evaluated steroids; six on bronchiolitis, two on Pneumocystis jirovecii (PJP) pneumonia, and one on croup.

#### Steroids for Bronchiolitis

Six RCTs evaluated steroids for bronchiolitis, four of which found no benefit for steroids on hospital LOS, duration of symptoms or Respiratory Distress Assessment Instrument (RDAI) score^51-54^. However, Teeratakulpisarn et al. (Thailand) found decreased time to resolution of respiratory distress, duration of symptoms, duration of oxygen therapy, and hospital LOS in children treated with single IM dexamethasone, and Virk et al. (Pakistan) found decreased rhonchi on day 3 of admission and shorter hospital LOS with oral prednisolone, although this study had high concern for bias.

#### Steroids for Croup, PJP pneumonia

Sumboonnanonda et al. (Thailand) found children treated with parenteral dexamethasone treatment had lower croup scores at 48 hours, shorter LOS and lower incidence of endotracheal intubation compared to placebo. Green et al. (South Africa) and Newberry et al. (Malawi), found that children with HIV and PJP pneumonia treated with adjunctive steroids had lower mortality rates.

### Medications – Other

Dawood et al. found no effect for oseltamivir in children with influenza on LOS or work of breathing in Panama. Adinatha (Indonesia) et al. and Becina Paolo Gene et al. (Philippines) found no benefit for adjuvant probiotics in children with pneumonia. Shang et al. (China) found children with bronchiolitis treated with *Laggera pterodonta* (Traditional Chinese Medicine herb), more frequently met discharge criteria at 96 and 120 hours compared to control group.

## DISCUSSION

Pediatric SARIs constitute a large burden of disease globally, with children in LMICs having the highest risk of mortality. Given the known association between delay of acute interventions with poor outcomes, we aimed to examine the current state of evidence on which emergency care interventions (those delivered in early period of care), have the strongest evidence for benefit on clinical outcomes. Patients generally present to a clinical provider without a specific disease or microbiologic diagnosis. Thus, our use of the SARI criteria as screening criteria was intentional in order to capture all studies including patients with undifferentiated respiratory illness. This strategy resulted in a broad array of interventions being included in the review, with most studies focused on medication interventions for children with pneumonia/ALRTIs or bronchiolitis (the most common respiratory conditions leading to hospitalization in children globally). Few interventions had strong evidence for benefit. The strongest evidence of benefit was found for respiratory support interventions. There was also strong evidence for lack of benefit of early use of adjuvant treatments such as Vitamin A, D and zinc on key clinical outcomes such as mortality and hospital length-of-stay for pneumonia and bronchiolitis.

### Respiratory Support Interventions including Bubble CPAP

The strongest evidence of benefit was found for respiratory support interventions. Strong oxygen delivery systems decrease risk of death, although results were inconclusive for the utility of CPAP specifically. Studies in this review show that respiratory support interventions should be implemented early, regardless of the final diagnosis: pneumonia, bronchiolitis, croup, other disease or undifferentiated. Particularly, interventions of equipment and supplies to deliver oxygen – concentrators, nasal devices and masks, must be readily available and combined with standardized training, technical support for maintenance of devices, pulse oximetry to monitor the patient, and guidelines for escalation and weaning of oxygen therapy. As a package, these interventions were found to decrease risk of death of a child with pneumonia by more than one third.

In general, CPAP was found to be very beneficial in children less than 1 year of age and somewhat beneficial in children less than 5 years of age with pneumonia in LMICs. However, this intervention should be used with caution as it can be particularly cumbersome for small children and requires extra financial, material and human resources to maintain. A 2021 systematic review evaluated CPAP for children with respiratory distress in resource-limited settings and concluded that the existing evidence is inconclusive for CPAP efficacy against death and adverse events, compared with oxygen^55^. The authors also point out the variations in resource availability, level of care available (ICU vs ward), and illness severity make the context in which interventions were assessed important for extrapolating results.

### Steroids and Other Treatments for Bronchiolitis

Steroids were not shown to be beneficial for bronchiolitis based on the findings in this review, but are beneficial for patients with HIV and PJP pneumonia, confirming earlier studies reflecting the same conclusions, including several systematic reviews finding a lack of convincing evidence for β2 agonists and anticholinergics, adrenaline, corticosteroids, hypertonic saline, antibiotics, and chest physiotherapy in the acute management of bronchiolitis, which has been reflected in current clinical practice guidelines^56^.

International treatment guidelines do not recommend nebulized saline for bronchiolitis, though a recent Cochrane review of articles from high- And low-income countries reported potentially improved outcomes (hospital LOS, clinical severity) based on low to moderate quality evidence^57^. In our review, one study with low risk of bias indicating nebulized NS may be beneficial for children with bronchiolitis^58^. However, the remaining eight studies that evaluated nebulized saline (either normal saline or hypertonic) with various comparators showed either no significant clinical outcome improvement ^59^ or had a high risk of bias associated with the study^60^.

In this review we found articles reporting mixed directionality of support for nebulized salbutamol for bronchiolitis, but unidirectional support in articles comparing benefit of nebulized epinephrine for bronchiolitis and croup. Though there were concerns of bias in some of the studies, this intervention should be considered as a targeted question for future international guideline updates.

### Early Administration of Adjuvant Nutrients and Minerals

Vitamin A only proved to be beneficial for children with measles and vitamin D only helpful for preventing repeat pneumonia^23,33^. A dedicated review of the vitamin D intervention for ALRTI international guideline development could be considered. Vitamin A, C or D use in other etiologies of SARI were not supported by the evidence, whether due to negative studies or risk of bias.

The two studies that evaluated zinc for bronchiolitis both had high risk of bias associated with them. While Farhad et al. found zinc to provide some clinical improvement, this has not been replicated, and zinc is not a recommended intervention for bronchiolitis by WHO guidelines^61^. However, among the sixteen articles that evaluated zinc for pneumonia, only one had high risk of bias associated. While most articles did not report significant clinical improvement in patients with pneumonia receiving zinc, and it is not a recommended standard intervention directly for pneumonia, zinc is an important micronutrient for a child’s overall health and development and is often deficient in children who are dehydrated^62,63^. It is plausible that for the studies in which zinc was found to have an improvement in clinical outcomes, it may have been because children in those studies were more dehydrated than in others. Zinc remains an important acute care intervention for children who are deficient or at risk of dehydration, but not part of regular home or inpatient treatment of children with pneumonia.

### Antibiotics

Antibiotics for pneumonia continue to be a recommended treatment according to international guidelines^61,64^. Differentiation is made between diagnoses and treatments for pneumonia and severe pneumonia. Pneumonia is a form of ALRTI and treated with oral amoxicillin as outpatient unless the child has HIV infection. Severe pneumonia is recommended to be treated initially with intravenous (IV) or intramuscular (IM) ampicillin (or benzylpenicillin) and gentamicin, switched to IV/IM gentamicin and cloxacillin if no improvement in 48 hours, and ceftriaxone in the case of failure of first-line treatment^61,64,65^. All three studies evaluating antibiotic regimens for severe pneumonia in children without underlying complications found treatment with high-dose oral amoxicillin equivalent to IV/IM ampicillin and recommended an update to current WHO guidelines. All articles were deemed to have some risk of bias, but this similar finding in these studies was replicated in nine countries across the three studies. We agree that this recommendation should be re-evaluated in international guidelines. Replacing parenteral antibiotics with an oral regimen for severe pneumonia would save time and money from hospitalization, including risk reduction of nosocomial infections. Ribeiro et al. showed that in comparison of IV oxacillin plus ceftriaxone versus of amoxicillin plus clavulanic acid for children (initiated IV and changed to oral after 48 hours) with very severe pneumonia, with outcomes showing significant improvement in tachypnea and decreased length of hospital stay in the amoxicillin group.

Chloramphenicol does not appear to be a viable option for severe pneumonia treatment based on the studies evaluated in this review and is not generally included as part of usual care for this illness^61^. Studies evaluating different antibiotic regimens for non-severe pneumonia revealed that cotrimoxazole is not a reliable first-line choice, and gentamicin is not required as a first line for community acquired pneumonia. Other regimens did not reveal a superior choice.

Multiple studies confirmed a lack of benefit antibiotics for treatment of viral-mediated illness (including bronchiolitis) supporting recommendations that antibiotics should not be used for viral respiratory infection. Gamino-Arroyo’s work evaluating nitazoxanide in addition to standard of care in children over 1 year of age in Mexico for influenza-like illness, as well as Kabir et al. and Pinto et al.’s work on evaluating antibiotics for viral bronchiolitis, are important additions to the literature supporting lack of effectiveness of antibiotics against viral illnesses, both in terms of lack of symptom improvement and in viral dynamics.

### Critical Gaps in the Literature

Very few studies included populations with undifferentiated respiratory distress, and only one study used the case definition of SARI as its inclusion criteria. Given that EC interventions often must be implemented prior to a formal clinical diagnosis, this review highlights a critical need for future studies to enroll children with undifferentiated respiratory infections. The majority (>80%) of the included studies evaluated medication interventions, with relatively few studies on other interventions, such as respiratory support. We also found that very few studies explicitly reported on the timing of interventions, with most studies either conducted outside of dedicated emergency wards or clearly during the earliest periods of care when the patient’s respiratory illness may be most amenable to acute interventions (e.g., shortly after presentation to a health facility, such as in triage or a dedicated emergency unit).

This review has several limitations. Limitations related to the development of the review and overall characteristics of results are previously described^7^. In particular and to emphasize, given that emergency care is not well defined across the world especially in LMICs and emergency care is often delivered outside of a dedicated emergency unit, we agreed on including any “emergent and early intervention” deemed appropriate by the reviewers, specifically including those focused on community-acquired pneumonia or discussing interventions beginning in the first 24 hours of hospitalization. As timing of interventions may greatly impact the trajectory of disease progression, future research on early and emergency care of patients must report timing and location of intervention in publications. Furthermore, inclusion of specific elements in the search strategy such as “bronchiolitis”, “croup” or “children” may have supported a smaller search result and tailored review.

## CONCLUSION

Despite few studies specifically evaluating patients in dedicated emergency units, or those with undifferentiated SARIs, a wide variety of interventions that may impact the early care of children was found. While the burden of SARI in pediatric populations is high, few interventions were found to have high quality evidence for benefit on clinical outcomes in LMICs, with respiratory support interventions having the strongest evidence for decreasing symptom duration and hospital length of stay, although with inconclusive evidence for bubble CPAP. Despite relatively few studies focused on non-medication interventions, the strongest evidence of benefit was found for respiratory support interventions such as improved oxygen delivery systems to decrease risk of death. Mixed results were found for interventions for bronchiolitis, although possible benefit was found for hypertonic nebulized saline to decrease hospital length of stay. Early use of adjuvant treatments such as Vitamin A, D and zinc did not appear to have convincing evidence of benefit on clinical outcomes in pneumonia and bronchiolitis. Further research evaluating the effect of targeted and time sensitive interventions for children with undifferentiated severe respiratory infections is greatly needed.

## Data Availability

All relevant data are within the manuscript and its Supporting Information files.

NA

## ACKNOWLEDGEMENTS

We thank Kelsey Sawyer of the Brown University Library for her assistance with development of the search strategies and retrieval. We thank the Global Emergency Medicine Literature Review (GEMLR) group and its editorial board for its support and dedication to this research.

## REFERENCES

1. Acute respiratory infections. 1990.

2. Evaluation IfHMa. Global Burden of Disease. 2019.

3. Orloff KE, Turner DA, Rehder KJ. The Current State of Pediatric Acute Respiratory Distress Syndrome. Pediatr Allergy Immunol Pulmonol. Jun 01 2019;32(2):35–44. doi:10.1089/ped.2019.0999

4. Jamison DT, Breman JG, Measham AR, et al. Disease Control Priorities in Developing Countries. 2006.

5. Fitzner J, Qasmieh S, Mounts AW, et al. Revision of clinical case definitions: influenza-like illness and severe acute respiratory infection. Bull World Health Organ. Feb 01 2018;96(2):122–128. doi:10.2471/BLT.17.194514

6. Global Emergency Medicine Literature Review Group.

7. Garbern SC, Relan P, O’Reilly GM, et al. A systematic review of acute and emergency care interventions for adolescents and adults with severe acute respiratory infections including COVID-19 in low- and middle-income countries. J Glob Health. Nov 08 2022;12:05039. doi:10.7189/jogh.12.05039

8. McGuinness LA, Higgins JPT. Risk-of-bias VISualization (robvis): An R package and Shiny web app for visualizing risk-of-bias assessments. Res Synth Methods. Jan 2021;12(1):55–61. doi:10.1002/jrsm.1411

9. Abdelbasset W, Elnegamy T. Effect of chest physical therapy on pediatrics hospitalized with pneumonia. International Journal of Health and Rehabilitation Science. 2015;4(4):219–226.

10. Lukrafka JL, Fuchs SC, Fischer GB, Flores JA, Fachel JM, Castro-Rodriguez JA. Chest physiotherapy in paediatric patients hospitalised with community-acquired pneumonia: a randomised clinical trial. Archives of Disease in Childhood. 2012;97(11):967–971.

11. Bohe L, Ferrero ME, Cuestas E, Polliotto L, Genoff M. Indicacion de la fisioterapia respiratoria convencional en la bronquiolitis aguda. Medicina (BAires). 2004;64(3):198–200.

12. Gomes ÉLFD, Postiaux G, Medeiros DRL, Monteiro KKDS, Sampaio LMM, Costa D. Chest physical therapy is effective in reducing the clinical score in bronchiolitis: randomized controlled trial. Braz j phys ther (Impr). 2012;16(3):241–247.

13. Singh M, Singhi S, Walia BN. Evaluation of steam therapy in acute lower respiratory tract infections: a pilot study. Indian Pediatr. 1990;

14. Addo-Yobo E, Chisaka N, Hassan M, et al. Oral amoxicillin versus injectable penicillin for severe pneumonia in children aged 3 to 59 months: a randomised multicentre equivalency study. Lancet. 2004;364(9440):1141–8. doi:10.1016/s0140-6736(04)17100-6

15. Hazir T, Fox LM, Nisar YB, et al. Ambulatory short-course high-dose oral amoxicillin for treatment of severe pneumonia in children: a randomised equivalency trial. Lancet. 2008;371(9606):49–56. doi:10.1016/s0140-6736(08)60071-9

16. Jeena P, Thea DM, MacLeod WB, et al. Failure of standard antimicrobial therapy in children aged 3-59 months with mild or asymptomatic HIV infection and severe pneumonia. Bull World Health Organ. 2006;84(4):269–75. doi:10.2471/blt.04.015222

17. Agweyu A, Gathara D, Oliwa J, et al. Oral amoxicillin versus benzyl penicillin for severe pneumonia among kenyan children: a pragmatic randomized controlled noninferiority trial. Clin Infect Dis. 2015;60(8):1216–24. doi:10.1093/cid/ciu1166

18. Ojha AR, Mathema S, Sah S, Aryal UR. A comparative study on use of 3% saline versus 0.9% saline nebulization in children with bronchiolitis. J Nepal Health Res Counc. 2014;12(26):39–43.

19. Sharma BS, Gupta MK, Rafik SP. Hypertonic (3%) saline vs 0.9% saline nebulization for acute viral bronchiolitis: a randomized controlled trial. Indian pediatrics. 2013;50(8):743–747.

20. Tinsa F, Abdelkafi S, Bel Haj I, et al. A randomized, controlled trial of nebulized 5% hypertonic saline and mixed 5% hypertonic saline with epinephrine in bronchiolitis. Tunis Med. 2014;92(11):674–7.

21. Si NV, Grytter C, Vy NN, Hue NB, Pedersen FK. High dose vitamin A supplementation in the course of pneumonia in Vietnamese children. Acta Paediatr. 1997;86(10):1052–5. doi:10.1111/j.1651-2227.1997.tb14805.x

22. Fawzi WW, Mbise RL, Fataki MR, et al. Vitamin A supplementation and severity of pneumonia in children admitted to the hospital in Dar es Salaam, Tanzania. Am J Clin Nutr. 1998;68(1):187–92. doi:10.1093/ajcn/68.1.187

23. Hussey GD, Klein M. A randomized, controlled trial of vitamin A in children with severe measles. N Engl J Med. 1990;323(3):160–4. doi:10.1056/nejm199007193230304

24. Nacul LC, Kirkwood BR, Arthur P, Morris SS, Magalhães M, Fink MC. Randomised, double blind, placebo controlled clinical trial of efficacy of vitamin A treatment in non-measles childhood pneumonia. Bmj. 1997;315(7107):505–10. doi:10.1136/bmj.315.7107.505

25. Rodríguez A, Hamer DH, Rivera J, et al. Effects of moderate doses of vitamin A as an adjunct to the treatment of pneumonia in underweight and normal-weight children: a randomized, double-blind, placebo-controlled trial. Am J Clin Nutr. 2005;82(5):1090–6. doi:10.1093/ajcn/82.5.1090

26. Stephensen CB, Franchi LM, Hernandez H, Campos M, Gilman RH, Alvarez JO. Adverse effects of high-dose vitamin A supplements in children hospitalized with pneumonia. Pediatrics. 1998;101(5):E3. doi:10.1542/peds.101.5.e3

27. Mahalanabis D, Jana S, Shaikh S, et al. Vitamin E and vitamin C supplementation does not improve the clinical course of measles with pneumonia in children: a controlled trial. Oxford; UK: Oxford University Press; 2006. p. 302–303.

28. Somnath SH, Biswal N, Chandrasekaran V, Jagadisan B, Bobby Z. Therapeutic effect of vitamin D in acute lower respiratory infection: A randomized controlled trial. Clin Nutr ESPEN. 2017;20:24–28. doi:10.1016/j.clnesp.2017.02.003

29. Choudhary N, Gupta P. Vitamin D supplementation for severe pneumonia--a randomized controlled trial. Indian Pediatr. 2012;49(6):449–54. doi:10.1007/s13312-012-0073-x

30. Gupta P, Dewan P, Shah D, et al. Vitamin D Supplementation for Treatment and Prevention of Pneumonia in Under-five Children: A Randomized Double-blind Placebo Controlled Trial. Indian Pediatr. 2016;

31. Rajshekhar CS, Vanaki R, Badakali AV, Pol RR, Yelamali BC. Efficacy of vitamin D supplementation in the treatment of severe pneumonia in children aged less than five years. Int J Contemp Pediatr. 2016;3(1):96–99.

32. Dhungel A, Alam MS. Efficacy of vitamin D in children with pneumonia: a randomized control trial study. 2016;3

33. Manaseki-Holland S, Qader G, Isaq Masher M, et al. Effects of vitamin D supplementation to children diagnosed with pneumonia in Kabul: a randomised controlled trial. Trop Med Int Health. 2010;15(10):1148–55. doi:10.1111/j.1365-3156.2010.02578.x

34. Ahadi A, Mirzarahimi M, Barak M, Ahari SS, Reyhanian M. Comparing the effect of zinc gluconate and placebo in the treatment of tachypnea, dyspnea and fever in children aged 2 to 23 months with acute bronchiolitis. 2020;7

35. Farhad H, Fatemeh B, Mohammadkhaje D, Hamidreza K, Mohammadnasir H. <The> role of zinc sulfate in acute bronchiolitis in patients aged 2 to 23 months. 2011. p. 231–234.

36. Bose A, Coles CL Gunavathi, et al. Efficacy of zinc in the treatment of severe pneumonia in hospitalized children <2 y old. Am J Clin Nutr. 2006;83(5):1089–96; quiz 1207. doi:10.1093/ajcn/83.5.1089

37. Brooks WA, Yunus M, Santosham M, et al. Zinc for severe pneumonia in very young children: double-blind placebo-controlled trial. Lancet. 2004;363(9422):1683–8. doi:10.1016/s0140-6736(04)16252-1

38. Coles CL, Bose A, Moses PD, et al. Infectious etiology modifies the treatment effect of zinc in severe pneumonia. Am J Clin Nutr. 2007;86(2):397–403. doi:10.1093/ajcn/86.2.397

39. Howie S, Bottomley C, Chimah O, et al. Zinc as an adjunct therapy in the management of severe pneumonia among Gambian children: randomized controlled trial. J Glob Health. 2018;8(1):010418. doi:10.7189/jogh.08.010418

40. Fataki MR, Kisenge RR, Sudfeld CR, et al. Effect of zinc supplementation on duration of hospitalization in Tanzanian children presenting with acute pneumonia. J Trop Pediatr. 2014;60(2):104–11. doi:10.1093/tropej/fmt089

41. Heydarian F, Nasiri M, Nakhaei AA, Ahanchian H, Ghahremani S, Haghbin A. Investigating the effect of prescribing zinc sulfate on improving the clinical symptoms of pneumonia in 2-59-month-old children. International Journal of Pediatrics. 2020;8(11):12471–12479.

42. Laghari GS, Hussain Z, Taimur M, Jamil N. Therapeutic Role of Zinc Supplementation in Children Hospitalized with Pneumonia. Cureus. 2019;11(4):e4475. doi:10.7759/cureus.4475

43. Mahalanabis D, Chowdhury A, Jana S, et al. Zinc supplementation as adjunct therapy in children with measles accompanied by pneumonia: a double-blind, randomized controlled trial. Am J Clin Nutr. 2002;76(3):604–7. doi:10.1093/ajcn/76.3.604

44. Qasemzadeh MJ, Fathi M, Tashvighi M, et al. The effect of adjuvant zinc therapy on recovery from pneumonia in hospitalized children: a double-blind randomized controlled trial. Scientifica (Cairo). 2014;2014:694193. doi:10.1155/2014/694193

45. Rerksuppaphol L, Rerksuppaphol S. Efficacy of Adjunctive Zinc in Improving the Treatment Outcomes in Hospitalized Children with Pneumonia: A Randomized Controlled Trial. J Trop Pediatr. 2020;66(4):419–427. doi:10.1093/tropej/fmz082

46. Semp√©rtegui F, Estrella B, Rodr√≠guez O, et al. Zinc as an adjunct to the treatment of severe pneumonia in Ecuadorian children: a randomized controlled trial. Am J Clin Nutr. 2014;99(3):497–505. doi:10.3945/ajcn.113.067892

47. Shah GS, Dutta AK, Shah D, Mishra OP. Role of zinc in severe pneumonia: a randomized double bind placebo controlled study. Ital J Pediatr. 2012;38:36. doi:10.1186/1824-7288-38-36

48. Srinivasan MG, Ndeezi G, Mboijana CK, et al. Zinc adjunct therapy reduces case fatality in severe childhood pneumonia: a randomized double blind placebo-controlled trial. BMC Med. 2012;10:14. doi:10.1186/1741-7015-10-14

49. Wadhwa N, Chandran A, Aneja S, et al. Efficacy of zinc given as an adjunct in the treatment of severe and very severe pneumonia in hospitalized children 2-24 mo of age: a randomized, double-blind, placebo-controlled trial. Am J Clin Nutr. 2013;97(6):1387–94. doi:10.3945/ajcn.112.052951

50. Valentiner-Branth P, Shrestha PS, Chandyo RK, et al. A randomized controlled trial of the effect of zinc as adjuvant therapy in children 2-35 mo of age with severe or nonsevere pneumonia in Bhaktapur, Nepal. Am J Clin Nutr. 2010;91(6):1667–74. doi:10.3945/ajcn.2009.28907

51. Aamer Q, Humayun K, Afsheen M. Comparison of mean duration of bronchiolitis in children receiving standard treatment with and without intravenous steroid. 2015. p. 5–9.

52. Ahmad Omair V. Comparison of conventional treatment along with prednisolone and conventional treatment alone in bronchiolitis. 2014. p. 160–163.

53. Mesquita M, Castro-Rodríguez JA, Heinichen L, Fariña E, Iramain R. Single oral dose of dexamethasone in outpatients with bronchiolitis: a placebo controlled trial. Allergol Immunopathol (Madr). 2009;37(2):63–7. doi:10.1016/s0301-0546(09)71106-1

54. Zhang L, Ferruzzi E, Bonfanti T, et al. Long and short-term effect of prednisolone in hospitalized infants with acute bronchiolitis. J Paediatr Child Health. 2003;39(7):548–51. doi:10.1046/j.1440-1754.2003.00212.x

55. Sessions KL, Smith AG, Holmberg PJ, et al. Continuous positive airway pressure for children in resource-limited settings, effect on mortality and adverse events: systematic review and meta-analysis. Arch Dis Child. Jun 2022;107(6):543–552. doi:10.1136/archdischild-2021-323041

56. Hartling L, Fernandes RM, Bialy L, et al. Steroids and bronchodilators for acute bronchiolitis in the first two years of life: systematic review and meta-analysis. BMJ. Apr 06 2011;342:d1714. doi:10.1136/bmj.d1714

57. Zhang L, Mendoza-Sassi RA, Wainwright C, Klassen TP. Nebulised hypertonic saline solution for acute bronchiolitis in infants. Cochrane Database Syst Rev. 12 21 2017;12:CD006458. doi:10.1002/14651858.CD006458.pub4

58. Pukai G, Duke T. Nebulised normal saline in moderate acute bronchiolitis and pneumonia in a low-to middle-income country: a randomised trial in Papua New Guinea. Paediatr Int Child Health. 2020;40(3):171–176. doi:10.1080/20469047.2020.1725338

59. !!! INVALID CITATION !!! 32–38;

60. A PK, Rajarathinam I, Gowdra A. Comparative evaluation of nebulised 3% saline versus nebulised 0.9% saline in the treatment of acute bronchiolitis. 2019;6

61. Pocket Book of Hospital Care for Children: Guidelines for the Management of Common Childhood Illnesses. 2013.

62. Wapnir RA. Zinc deficiency, malnutrition and the gastrointestinal tract. J Nutr. May 2000;130(5S Suppl):1388S–92S. doi:10.1093/jn/130.5.1388S

63. Roy SK, Tomkins AM, Ara G, et al. Impact of zinc deficiency on vibrio cholerae enterotoxin-stimulated water and electrolyte transport in animal model. J Health Popul Nutr. Mar 2006;24(1):42–7.

64. UNICEF. Standard Treatment Manual for Children: A manual for health workers. 4th ed. 2017.

65. Tong N. Background Paper 6.22 Pneumonia: Update on 2004 Background Paper. Priority Medicines for Europe and the World “A Public Health Approach to Innovation”: World Health Organization; 2013.

